# Scalable estimation of temporal clustering in accelerometry: a kernel-independent dispersion index grounded in the Hawkes process

**DOI:** 10.64898/2026.06.14.26355611

**Authors:** Xin Zheng, Ian Meneghel Danilevicz, Maichina Paw

**Affiliations:** Amsterdam UMC location Vrije Universiteit Amsterdam, Public and Occupational Health, De Boelelaan 1117, Amsterdam, the Netherlands; Amsterdam Public Health, Health Behaviors and Chronic Diseases and Methodology, Amsterdam, the Netherlands; Université Paris Cité, Inserm U1153, Centre for Research in Epidemiology and Statistics (CRESS), Epidemiology of Ageing and Neurodegenerative Diseases (EpiAgeing), Paris, France

**Keywords:** dispersion index, Hawkes process, branching ratio, temporal clustering, point process estimation, accelerometry, mortality

## Abstract

**Background:** Self-exciting (Hawkes) point processes are a natural model for the temporal clustering of human physical activity (PA) recorded by accelerometers, yet they have seldom been used in this setting—in part because the usual maximum-likelihood fitting is challenging due to potential estimation bias and convergence failures on these data. A moment-based alternative—estimating the Hawkes branching ratio from the dispersion index, the variance-to-mean ratio of event counts—is kernel-independent and computationally trivial, but it has not been evaluated for accelerometry or adapted to the intensity-marked recordings accelerometers provide.

**Methods:** Treating each minute above a sedentary threshold as an event, we estimated the Hawkes branching ratio *n* by maximum likelihood and, as a kernel-independent and far cheaper alternative, from the dispersion index. We compared four dispersion-based estimators—event-count-based, intensity-mark-weighted using the mark-moment ratio, and time-of-day (TOD) adjusted variants of each—against the marked and unmarked maximum-likelihood estimates. Estimators were evaluated for mutual agreement, goodness of fit, and finite-window results in two National Health and Nutrition Examination Survey (NHANES) accelerometry cohorts (hip-worn, *n* = 2,560; wrist-worn, *n* = 3,132). We related the resulting temporal clustering measures to all-cause mortality using survey-weighted Cox models, adjusting for PA frequency, Peak30 (the average of the 30 highest PA values), and demographic covariates.

**Results:** Event-count-based dispersion estimates agreed strongly with maximum-likelihood branching ratios (*r* ≈ 0.74 in both cohorts); the intensity-marked variant incorporating PA intensity variability agreed less well. Marked and unmarked Hawkes models yielded similar excitation and decay parameters, suggesting PA intensity added little clustering information beyond event timing. In the survival analysis, temporal clustering was associated with all-cause mortality independently of PA frequency and Peak30; the direction of association differed between the hip- and wrist-worn cohorts.

**Conclusions:** A scalable dispersion-index estimator recovers the Hawkes branching ratio and matches maximum-likelihood estimates without requiring kernel specification or iterative optimization. It offers a practical tool for quantifying temporal clustering in accelerometry, enabling decomposition of temporal PA patterns into its exogenous initiation and endogenous persistence. Such temporal patterns carry health-relevant information beyond PA intensity and volume.

## 1 Background

Regular physical activity (PA) confers substantial health benefits, including reduced risks of cardiovascular disease, cancer, and all-cause mortality [1, 2, 3]. Yet the temporal patterns– how individuals accumulate PA throughout the day–vary widely across individuals. Whether such temporal patterns across different intensity levels carry additional health-relevant information beyond total PA volume remains an interesting question, which is challenging to answer [4, 5, 6].

A large and growing literature has sought to characterize temporal movement behavior from wearable-device data. In the most comprehensive review to date, Kobeissi *et al*. [7] identified 161 standalone metrics and methods. Their inventory shows that current approaches have focused primarily on *volume* (e.g. Peak 1/30/60-min PA values [8]), *intensity distribution* (e.g. average time spent at different intensity levels), *bout characteristics* (e.g. duration and frequency) [9, 10, 11], and variance-based indicators such as interdaily stability, intradaily variability, and the coefficient of variation (CoV) [12, 13, 14]. Several more advanced temporal summaries–spanning complexity, regularity and burstiness–have also been proposed, including the power-law exponent and Gini index for sedentary-bout distributions [15], sample entropy for time-series regularity [16], and fragmentation indices based on mean bout duration [17, 18, 19]. Although these metrics reorganize the same high-resolution accelerometry signal in different ways, they rarely model human behavioral dynamics as a *stochastic process*; detrended fluctuation analysis and the self-similarity parameter are among the few that do [20, 18].

Self-exciting point processes, and the Hawkes process [21] in particular, fill that gap. PA clustering manifests as temporal autocorrelation [22]: active epochs do not occur independently but tend to trigger further active epochs in close temporal proximity. The Hawkes framework captures this directly by treating active epochs as discrete events on continuous time and decomposing the resulting event sequence into a baseline (exogenous initiation) component and a self-exciting (endogenous persistence) component in which past epochs raise the probability of future ones [23]. Activity bouts then emerge from self-excitation rather than being imposed by a duration threshold. Originally developed for seismology [24], Hawkes processes have since been applied across finance, criminology, and infectious-disease modeling [25, 26, 27, 28]. Paraschiv-Ionescu *et al*. [29] first brought them to PA, using a bivariate model of activity-rest transitions in chronic-pain patients and showing that behavioral dynamics are embedded in temporal patterning. Their approach, however, required maximum-likelihood estimation (MLE) of several parameters, limiting its scalability to large epidemiological studies.

Hawkes MLE faces several well-documented difficulties [30] that also apply to PA data. Maximum-likelihood procedures tend to return branching ratiosthe expected number of events triggered by a single eventapproaching unity [31]. In this near-critical regime, external temporal trends are spuriously absorbed into the excitation mechanism, a likely failure mode for accelerometry given strong time-of-day (TOD) non-stationarity [32]; the fitted conditional intensity then tracks individual sequences through highly flexible trajectories, and parameter estimates become unstable across data perturbations and optimization initializations [31], eroding the physical interpretation of self-excitation. Crucially, near-critical fits often produce excellent goodness-of-fit (GOF) diagnostics, including near-perfect Kolmogorov– Smirnov (KS) agreement, so overfitting is hard to detect [33]; kernel misspecification can likewise yield small KS distances alongside badly biased estimates [32, 34]. Satisfactory GOF, in short, does not by itself certify model adequacy here. As a partial remedy, Hardiman & Bouchaud [35] showed that the branching ratio can be estimated directly from the dispersion index (the variance-to-mean ratio of event counts). The relationship is kernel-independent—it holds for any stationary Hawkes kernel, not only the exponential—and requires only the first two moments of windowed counts, which makes it attractive precisely when the excitation mechanism is unknown and MLE is fragile.

To our knowledge this estimator has not been evaluated on accelerometry, nor adapted to the marked Hawkes processes that intensity-marked PA data naturally motivate. Using large accelerometry datasets from NHANES 2003–2006 (hip-worn) and 2011–2014 (wrist-worn) [36, 37], we (1) evaluate whether the dispersion-index branching-ratio estimator is a scalable, kernel-independent alternative to Hawkes MLE, validating the two against each other; (2) extend the estimator to marked Hawkes processes and ask whether PA intensity modulates temporal clustering beyond event timing; and (3) relate the scalable temporal clustering measure to all-cause mortality, demonstrating its prognostic value beyond PA volume.

## 2 Theoretical framework

### 2.1 Active epochs as a point process

Let {*t*_*i*_} _*i*≥ 0_ denote the times of active epochs—minutes during which physical activity (PA) intensity exceeds the sedentary threshold. We model these as a temporal point process with associated counting measure *N* (*A*) = # {*i* : *t*_*i*_ ∈ *A*} and conditional intensity *λ*(*t*) [23]. Self-excitation in this framework captures the tendency of activity to sustain itself minute-by-minute, within and across bouts (sustained activity across many consecutive active epochs), producing overdispersion (high variance relative to mean) in event counts.

### 2.2 The Hawkes process

The Hawkes process [21] models self-exciting events through the conditional intensity

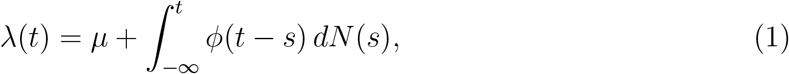

where *µ* > 0 is the baseline intensity, *ϕ*(*τ* ) ≥ 0 the excitation kernel, and 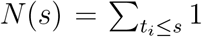 the counting process of {*t*_*i*_}. The *branching ratio*

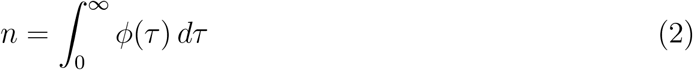

is the expected number of offspring events triggered by each parent event; stationarity requires *n* < 1. For the exponential kernel *ϕ*(*τ*) = *αe*^−*βτ*^ the conditional intensity becomes [38]

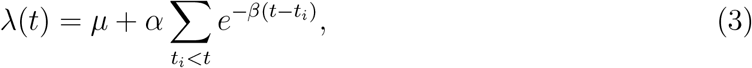

with *n* = *α/β*, where *α* is the amplitude of self-excitation and *β* its decay rate; under stationarity the expected intensity is Λ = *µ/*(1 − *n*).

### 2.3 The dispersion index and the branching ratio

The dispersion index (also called the variance-to-mean ratio or Fano factor [39]) for event counts in a window of size *W* is

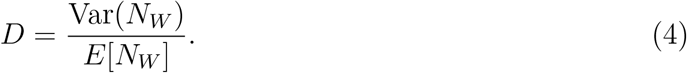

For a Poisson process *D* = 1; for a clustered (self-exciting) process *D* > 1. The following result links this elementary summary to the branching ratio.

#### Theorem 1

(Dispersion–branching relation, unmarked process). *For any stationary Hawkes process with branching ratio n* < 1, *the asymptotic (large-window) dispersion index of event counts is*

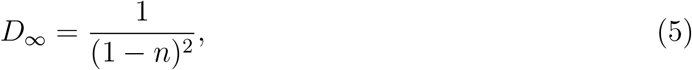

*independently of the kernel shape ϕ. Equivalently, the branching ratio is recovered as*

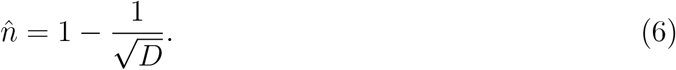

#### Proof

Hardiman & Bouchaud [35] obtained (5) via the spectral representation of the Hawkes process. We give the equivalent cluster-representation derivation [40], which is more directly interpretable. The Hawkes process admits a branching structure: events arriving from the baseline intensity *µ* act as immigrants, and each event—immigrant or descendant— independently triggers offspring according to a branching process with mean *n*. Let *C* denote the total size of a cluster (an immigrant plus all its descendants), defined as 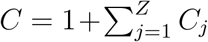, where *Z* ∼Poisson(*n*) is the number of direct offspring and *C*_*j*_ are i.i.d. copies of *C*. The random variable *C* is said to have a Borel distribution with parameter *n*, written *C* ∼ Borel(*n*) [41]. So standard branching-process theory gives

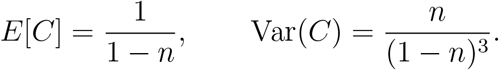

The total count in a large window of size *T* is 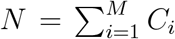, where *M* ∼ Poisson(*µT* ) is the number of immigrants and the *C*_*i*_ are independent copies of *C*. Since immigrants arrive as a Poisson process and generate independent clusters, *N* follows a compound Poisson distribution, so *E*[*N* ] = *E*[*M* ] *E*[*C*] and Var(*N* ) = *E*[*M* ] *E*[*C*^2^]. Using *E*[*C*^2^] = Var(*C*) + *E*[*C*]^2^ = (1 − *n*)^−3^,

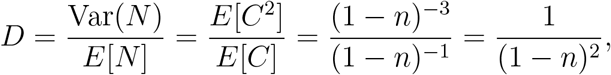

and inverting yields (6).

The estimator depends only on *n*, not on the kernel shape, and so holds for exponential, power-law, or any other kernel. The intuition is transparent: clustering inflates variance because some windows contain entire clusters (bursts) while others fall between them (silence). The derivation assumes windows large enough to contain complete clusters; for finite windows, clusters truncated at boundaries give *D*(*W* ) < *D*_∞_, so 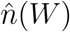 underestimates *n* and converges to the true value as *W* → ∞ [35].

#### 2.3.1 Time-of-day (TOD) adjustment

Human activity exhibits strong TOD variation due to circadian rhythms, violating the stationarity assumption. The event-count dispersion index then conflates this TOD variation with self-excitation-driven clustering and overestimates *n*. We address this by detrending each window with a TOD baseline, adapting to daily activity rhythms the intraday-seasonality adjustment Hardiman & Bouchaud [35] applied to financial data.

For a single day, the observation period is partitioned into non-overlapping windows of width *W* ; let ℐ_*j*_ be the index set of events in window *j* and 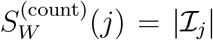 the corresponding event count. The raw event-count dispersion index and recovered branching ratio are

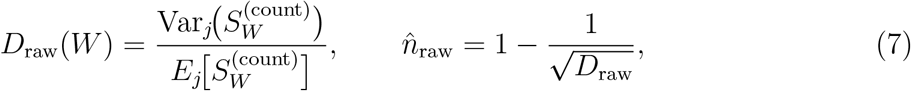

the finite-window counterpart of Theorem 1. To detrend, the day is partitioned into *H* TOD bins. With *N*_*h*_ the number of events in bin *h* and *H*^seg^ the day’s activity segment (the contiguous run of bins from the first to the last event), the TOD baseline is

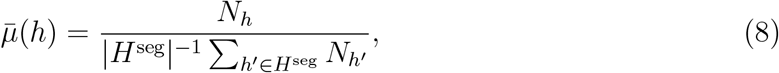

normalized to average one over the segment, so 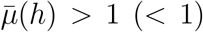 marks times of day with above-(below-) average activity. Because the bins are coarser than the counting windows, we apply a single shared factor per window—the average baseline over its events—

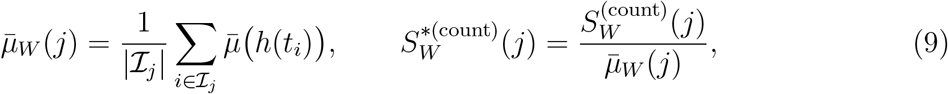

which downweights windows falling in habitually high-activity periods. The TOD-adjusted dispersion index and branching ratio are then

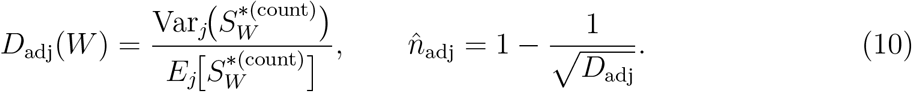

Detrending removes deterministic exogenous variation and isolates the endogenous clustering component; the adjusted index is reported alongside the unadjusted one.

### 2.4 Marked extension: does intensity modulate clustering?

Unmarked Hawkes processes treat all active epochs identically, yet accelerometry intensities are heavy-tailed [34]: each active epoch carries a mark *m*_*i*_ representing its activity intensity (counts or MIMS), from light to vigorous. To test whether self-excitation depends on intensity, we consider a marked model where each event’s excitatory influence scales with its intensity-mark (*m*_*i*_) [23],

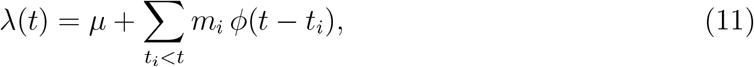

so that higher-intensity epochs would generate stronger self-excitation if intensity genuinely modulated clustering. The dispersion index then absorbs variance from both temporal clustering and intensity-mark variability, in a form that parallels Theorem 1.

#### Theorem 2

(Dispersion–branching relation, marked process). *For a stationary marked Hawkes process* (11) *with i*.*i*.*d. marks of finite second moment, the asymptotic dispersion index of the windowed mark sum is*

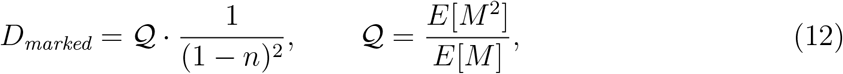

*where* 𝒬 *is the mark-moment ratio, and the branching ratio is recovered as*

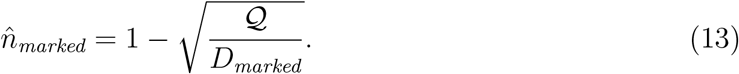

The full derivation is given in Supplementary Material I. It is the same compound-Poisson cluster argument used for Theorem 1, modified at a single step: because an event’s mark governs its offspring, the parent mark enters the cluster-mass moments. When marks are constant, 𝒬 reduces to that constant and (12) collapses to Theorem 1. The comparison of (13) with (6) is thus a direct test whether PA intensity modulated clustering.

Operationally, the marked estimator replaces the count with the windowed mark sum 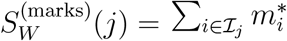 over the windows of Section 2.3.1 (intensity marks 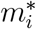 normalized to mean one per day), giving

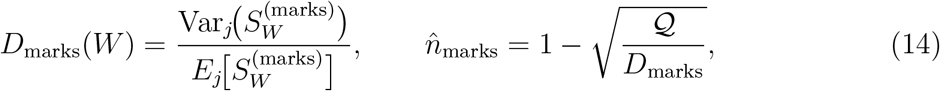

with 𝒬= *E*[*M* ^2^]*/E*[*M* ] estimated from the day’s marks. The TOD adjustment carries over directly: applying the same count-based baseline 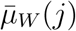 of (9) to the mark sum, 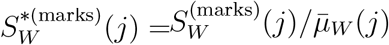, gives

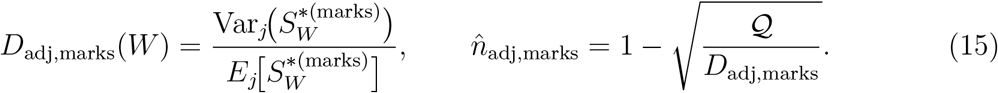

Under Theorem 2‘s assumption that marks are independent of event times, the event-count-based and intensity-mark-sum baselines coincide, and the shared normalization makes any gap between the unmarked and marked adjusted indices attributable to the marks rather than to a difference in detrending. Equations (7)–(15) define the four estimators carried forward to the empirical analysis: 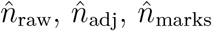, and 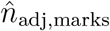. Daily dispersion indices were averaged across valid days to obtain participant-level estimates.

### 2.5 Behavioral interpretation

From the stationarity condition Λ = *µ/*(1 − *n*) we recover the exogenous initiation rate *µ*^∗^ = Λ^∗^(1 − *n*^∗^), where Λ^∗^ =∑_*i*_ *x*_*i,p*_*/T*_*p*_ is participant *p*’s empirical event frequency over valid observation time *T*_*p*_. For active epochs, *n*^∗^ captures the tendency of activity to sustain itself, while *µ*^∗^ captures how often new bouts are exogenously initiated; Table 1 summarizes the behavioral implications.

**Table 1:** Interpretation of key model parameters.

| Parameter | Mechanism | Level | Interpretation | Behavioral meaning |
| --- | --- | --- | --- | --- |
| $n^*$ | Endogenous (persistence) | Low | Fragmented events | Short activity bouts |
|  |  | High | Sustained bursts | Long activity bouts |
| $\mu^*$ | Exogenous (initiation) | Low | Infrequent initiation of events | Long sedentary intervals |
|  |  | High | Frequent initiation of events | Short sedentary intervals |

The decomposition into exogenous initiation (*µ*^∗^) and endogenous persistence (*n*^∗^) suggests potential intervention targets: individuals whose low activity levels are driven primarily by low *µ*^∗^ may benefit from prompts to initiate activity, whereas those whose low activity levels stems primarily from low *n*^∗^ may need support to sustain activity once started.

A further useful property is that the branching ratio *n* = *α/β* is time-scale invariant— rescaling all event times rescales *α* and *β* proportionally, leaving *n* unchanged—so estimates of temporal clustering are not confounded by measurement resolution (e.g., seconds vs. minutes), and reflect an intrinsic property of the activity process rather than an artifact of the time units.

## 3 Materials and methods

### 3.1 Study population

We analyzed accelerometry data from two National Health and Nutrition Examination Survey (NHANES) [42] cycles, which provide nationally representative samples of non-institutionalized U.S. adults aged 18–84.

#### Hip-worn accelerometry (2003–2006)

NHANES cycles C and D deployed ActiGraph 7164 accelerometers worn on the right hip for 7 consecutive days. Counts were recorded in 1-minute epochs during waking hours only.

#### Wrist-worn accelerometry (2011–2014)

NHANES cycles G and H used ActiGraph GT3X+ accelerometers worn on the non-dominant wrist for 7 consecutive days, recording tri-axial acceleration summarized in 1-minute epochs as Monitor-Independent Movement Summary (MIMS) units [37], with continuous 24-hour wear including sleep.

Covariates included demographics (age, sex, education, and race), health behaviors (smoking status, alcohol consumption), body mass index (BMI), cardiovascular risk factors (systolic blood pressure, total and high-density lipoprotein [HDL] cholesterol), and physician-diagnosed chronic conditions (diabetes, congestive heart failure, coronary heart disease, stroke, cancer). Mortality status and follow-up time were obtained from the NHANES-linked mortality files through 2019.

### 3.2 Data preprocessing

Preprocessing comprised quality control (Section 3.2.1) of the accelerometer data followed by event extraction for Hawkes modeling (Section 3.2.2); the participant flow is shown in Supplementary Figure 1. The hip-worn cohort began with 14,631 participants, of whom 2,560 were retained (17,920 person-days); the wrist-worn cohort began with 6,905 participants, of whom 3,132 were retained (21,924 person-days). Participant characteristics are given in Supplementary Table 1.

#### 3.2.1 Quality control

Quality control followed Leroux *et al*. [36] for the hip-worn data and was adapted to a comparable pipeline for the wrist-worn data: (1) wear/non-wear classification using the established NHANES algorithm [36]; (2) valid-day criteria requiring ≥ 10 hours of wake wear across 7 consecutive valid days; (3) retention of the longest continuous wear segment after removing interruptions exceeding 60 minutes; (4) sedentary thresholds of < 100 counts · min^−1^ for hip-worn data [36] and < 10.558 MIMS · min^−1^ for wrist-worn data [43, 44, 45]; and (5) exclusion of days with fewer than 5 active events.

#### 3.2.2 Event extraction

Because a stationary Hawkes process assumes time-invariant parameters—an assumption that TOD variation in PA may violate—we converted minute-level data into continuous event sequences through five steps: (1) events were defined as minutes exceeding the sedentary threshold; (2) bidirectional boundary trimming constrained each sequence to begin and end with a sedentary period, so that every sequence represented a complete self-exciting cycle; (3) quantization-noise correction via conservative temporal jittering mitigated the 1-minute discrete resolution of the raw data, restoring the continuous-time assumptions of the framework without altering event ordering; (4) event times were normalized to begin at *t* = 0 each day; and (5) marks were defined as PA values normalized (amplitude scaling) to mean 1 per day while preserving relative variation. Owing to the extreme positive skew and high variance of raw hip-worn counts, these were square-root transformed before modeling; the lower-variance MIMS units were not transformed.

### 3.3 Dispersion-index computation

We computed the four dispersion-based estimators defined in Sections 2.3.1 and 2.4—the event-count-based and intensity-mark-weighted indices and their TOD-adjusted variants— for each valid day. The observation period was divided into non-overlapping windows of width *W* ∈ {15, 30, 60, 90, 120} min (incomplete tail windows dropped so that retained windows had equal duration), and the TOD baseline used *H* = 12 two-hour bins; the mark-moment ratio 𝒬= *E*[*M* ^2^]*/E*[*M* ] was computed from each day’s marks. Daily indices and 𝒬 were averaged across valid days to the participant level. We used *W* = 30 min for the applied analysis, balancing finite-window truncation bias against the number of windows available per day for stable variance estimation; the simulation study (Section 4) justifies this choice.

### 3.4 Validation against Hawkes MLE

As a reference, we fitted stationary marked Hawkes models by MLE [38, 46]. For day *d*,

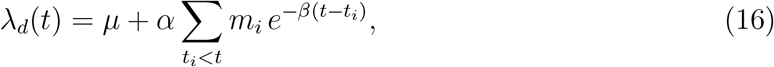

with branching ratio *n*_MLE_ = *E*(*m*) · *α/β*. With marks normalized to *E*[*M* ] = 1, the marked and unmarked branching ratios coincide and any divergence in fitted parameters would signal intensity-dependent self-excitation.

Having preprocessed each day toward stationarity (Section 3.2.2), we pooled the 7 days (weekly) data as independent realizations sharing (*µ, α, β*), rather than concatenating across midnight boundaries. The joint log-likelihood is

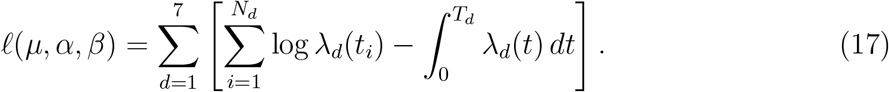

We then assessed agreement among the four dispersion estimators and the two MLE estimators.

Goodness of fit (GOF) used the random-time-change theorem [47]: under correct specification the compensator-transformed inter-arrivals 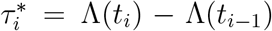 are i.i.d. Exp(1), checked via Q–Q plots, the Kolmogorov-Smirnov (KS), and Ljung–Box tests (Supplementary Material II). Treating each day as a separate realization, we computed 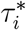 per day from each participant’s shared (*µ, α, β*) fit and assessed GOF at the per-day level.

### 3.5 Applied analysis: survival models

To assess whether the resulting clustering measures carries prognostic information, we used survey-weighted Cox proportional-hazards models for all-cause mortality, accounting for NHANES complex survey design with appropriate sampling weights and variance estimation. The primary exposures were the branching ratio *n* (temporal clustering) and the event rate *λ* (activity frequency); we used *λ* rather than *µ* because it directly represents the frequency dimension (events per unit time) and is less collinear with *n* than *µ*, which the stationarity relation *λ* = *µ/*(1 − *n*) constrains. Exposures were modeled per standard deviation (SD). Model 1 adjusted for age, sex, race, and wear time; Model 2 additionally adjusted for Peak30, BMI, smoking, drinking, education, mobility problems, and comorbidities (diabetes, cardiovascular disease, cancer, stroke).

## 4 Simulation study

### 4.1 Design

To validate the estimator against a known ground truth, we generated synthetic event sequences from three point processes with known properties, for *N* = 50 participants over 7 days each: (i) a homogeneous Poisson process (rate 20 events/hour): expected *D* ≈ 1,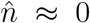; (ii) a self-exciting Hawkes process (exponential kernel, true branching ratio *n* = *α/β* = 0.6, decay *β* = 0.15 min^−1^, stationary rate 20 events/hour, baseline *µ* = *λ*(1 − *n*) = 8 events/hour): expected *D*_∞_ = 1*/*(1 − *n*)^2^ = 6.25, 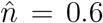; and (iii) a regular (underdispersed) process of evenly spaced events with ±10% jitter: expected *D* < 1, 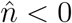.

Simulation parameters were chosen to approximate the empirical data: events were generated over a 14-hour window (08:00–22:00) at a rate of 20 events per hour, yielding approximately 280 events per participant-day, consistent with active-epoch rates observed in the NHANES data after preprocessing and event extraction. Hawkes processes were simulated via the thinning algorithm using the R package `emhawkes` [48]. Dispersion indices were computed at each candidate window *W* ∈ {5, 10, 15, 30, 45, 60, 90, 120} min. This validation targets the event-count-based estimator under a stationary, unmarked generative model—the regime in which Theorem 1 holds and the estimator we ultimately recommend; recovery under heavy-tailed marks or TOD non-stationarity is not simulated (see Discussion).

### 4.2 Window-size selection

The dispersion index correctly discriminated all three processes at every window size (Figure 1): *D* remained near unity for the Poisson process, well above unity for the Hawkes process, and below unity for the regular process. As expected by finite-window truncation (Theorem 1), the Hawkes 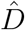 rose monotonically with *W* and so underestimated the asymptotic *D*_∞_ = 6.25; correspondingly, the recovered branching ratio increased from 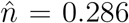 at *W* = 5 min to 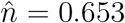 at *W* = 120 min, bracketing the true value *n* = 0.6. Estimation variance also grew with *W*, however, as fewer non-overlapping windows remained per day (e.g., 7 windows at *W* = 120 versus 28 at *W* = 30). We therefore adopted *W* = 30 min throughout, as a principled compromise: over the 840-minute observation period it yields 28 non-overlapping windows per day, providing sufficient samples for stable dispersion index estimation while maintaining temporal resolution below typical bout durations. This establishes that the event-count-based estimator recovers a known branching ratio before it is applied to free-living accelerometry data.

**Figure 1:**
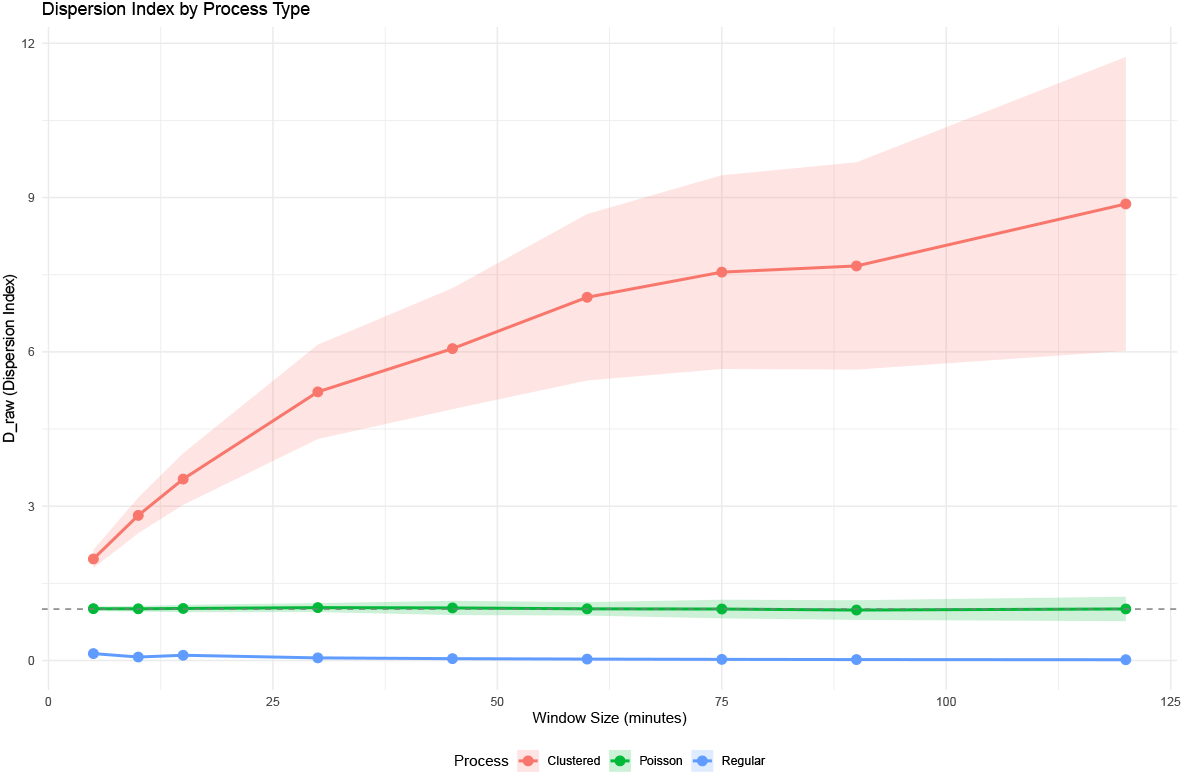
Validation of the dispersion index across window sizes for three simulated point processes: Poisson (homogeneous), Hawkes (self-exciting, *n* = 0.6, *β* = 0.15 min^−1^), and regular (underdispersed). Shaded ribbons denote ±1 SD across *N* = 50 participants over 7 days.

## 5 Results

### 5.1 Estimator in accelerometry across window sizes

Theorem 1 predicts that the raw dispersion index grows with window size due to finite-window truncation. The raw *D*_raw_ exhibited this expected growth with *W* (Figure 2), whereas the TOD-adjusted *D*_adj_ was approximately flat after removing the TOD trend. Both exceeded 1 for almost all participants, confirming persistent clustering beyond TOD effects for human PA behavior. The orderings *D*_marks_ > *D*_raw_ and *D*_adj,marks_ > *D*_adj_ held across window sizes, showing that intensity variation contributes additional dispersion; whether it carries additional *temporal clustering* information is the question taken up next.

**Figure 2:**
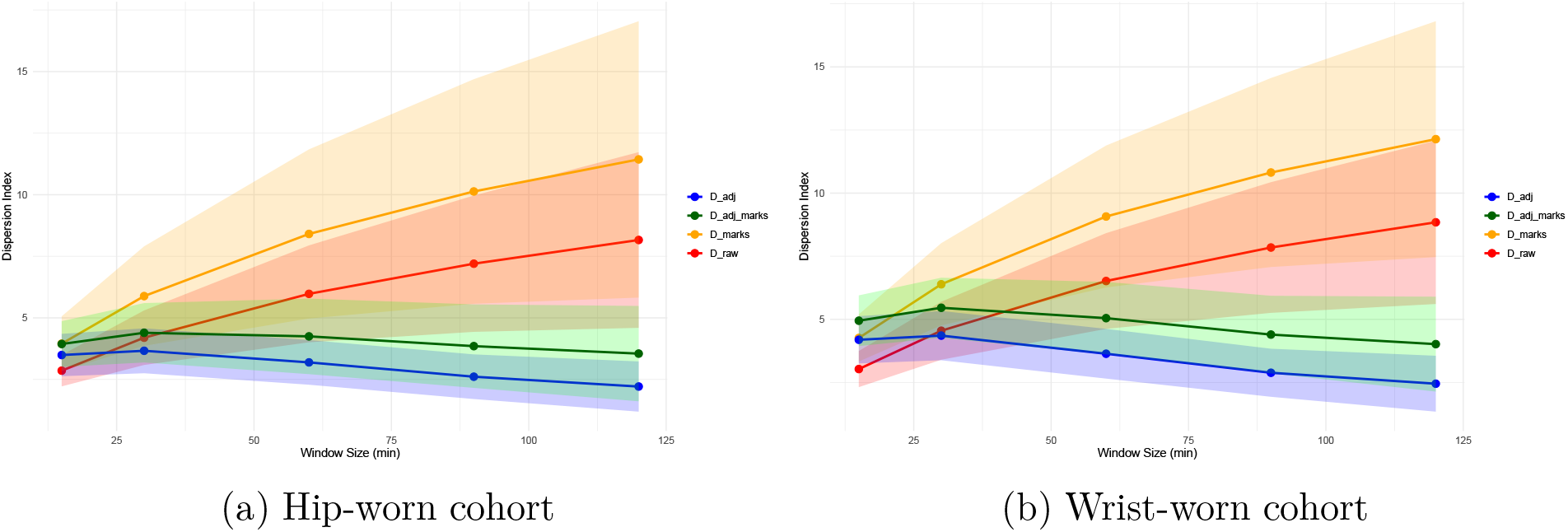
Dispersion index across window sizes. *D*_raw_ (event-count-based), *D*_adj_ (TOD-adjusted event-count), *D*_marks_ (intensity-mark-weighted), and *D*_adj_marks_ (TOD-adjusted intensity-mark).

### 5.2 Hawkes MLE reference fits and goodness of fit

We fitted marked and unmarked Hawkes models as the estimation reference. For the two illustrative individuals with weekly series in Figures 3a–3b (one per NHANES cohort, daily series in Supplementary Figures 2–17), branching ratio of about 0.75 indicated that roughly three-quarters of active epochs were endogenously triggered, and decay rates *β* ≈ 0.067– 0.082 min^−1^ corresponded to excitation timescales of about 12–15 min (1*/β*), so triggering dissipated within roughly a quarter of an hour. In the marked model (Table 2), the pooled means (hip 1.018; wrist 1.005) and SDs (hip 1.105; wrist 1.040) of the compensator-transformed inter-arrivals were close to the Exp(1) target, KS statistics stayed within acceptable limits (hip mean 0.100, max 0.150; wrist mean 0.127, max 0.137), Q–Q plots confirmed broad agreement, and all daily series passed Ljung–Box (*p* > 0.05).

**Table 2:**
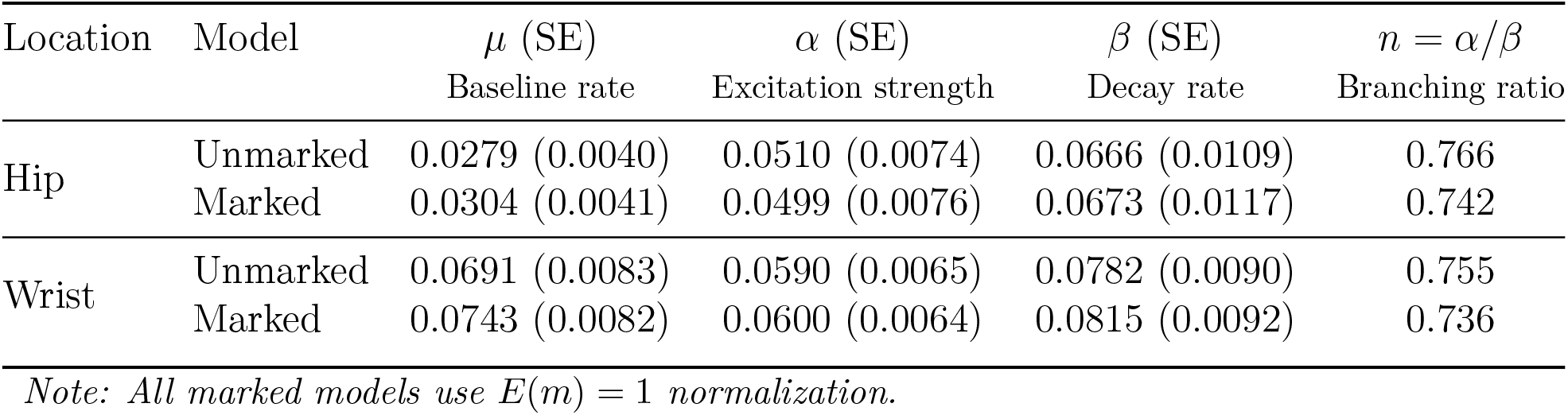
Comparison of parameter estimates for unmarked and marked Hawkes processes in the hip- and wrist-worn cohorts.

| Location | Model | $\mu$ (SE) | $\alpha$ (SE) | $\beta$ (SE) | $n = \alpha/\beta$ |
| --- | --- | --- | --- | --- | --- |
|  |  | Baseline rate | Excitation strength | Decay rate | Branching ratio |
| Hip | Unmarked | 0.0279 (0.0040) | 0.0510 (0.0074) | 0.0666 (0.0109) | 0.766 |
|  | Marked | 0.0304 (0.0041) | 0.0499 (0.0076) | 0.0673 (0.0117) | 0.742 |
| Wrist | Unmarked | 0.0691 (0.0083) | 0.0590 (0.0065) | 0.0782 (0.0090) | 0.755 |
|  | Marked | 0.0743 (0.0082) | 0.0600 (0.0064) | 0.0815 (0.0092) | 0.736 |

*Note: All marked models use $E(m) = 1$ normalization.*

**Figure 3:**
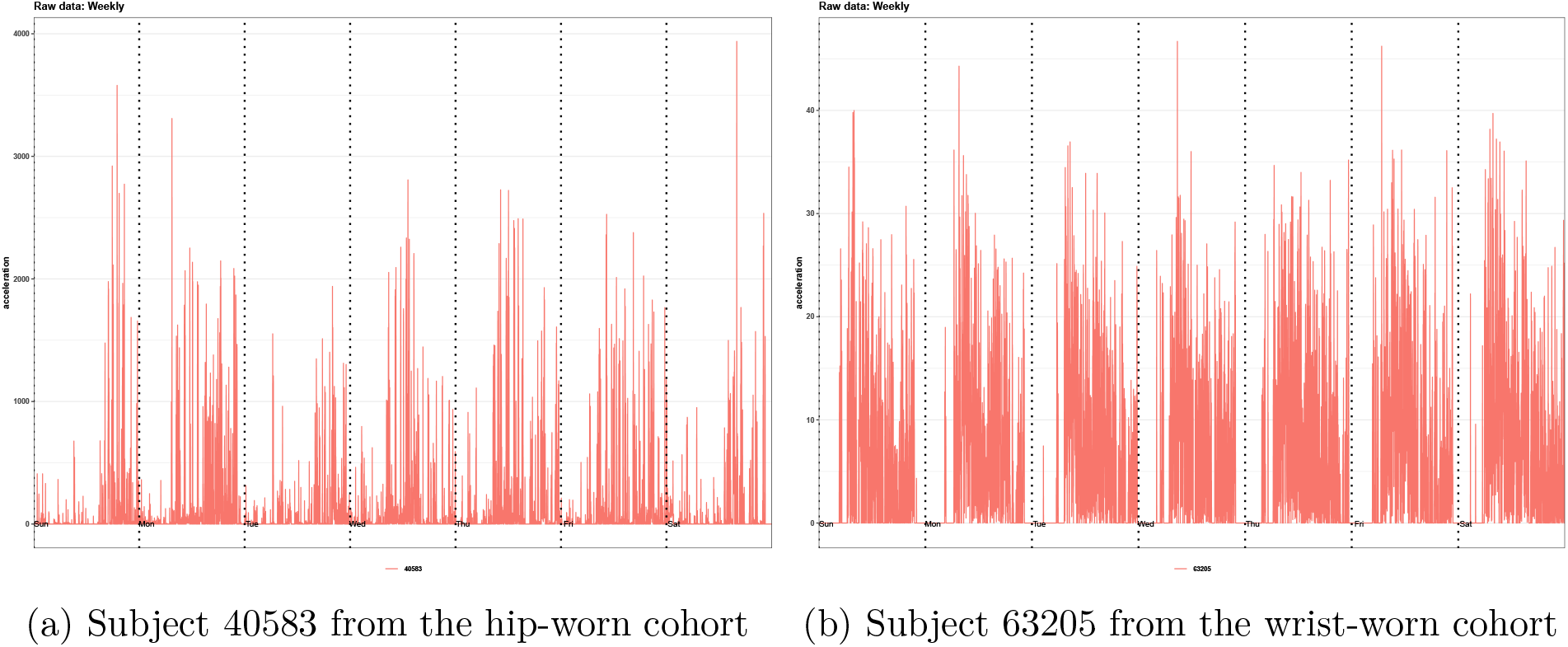
Illustrative weekly accelerometry data for one participant per cohort.

Across the full set of fitted participants, the marked and unmarked fits showed the same pattern—consistently high branching ratios and good residual agreement, with most daily series passing Ljung–Box (*p* > 0.05); the cross-participant distributions of these estimates are discussed in the next section.

### 5.3 Does intensity modulate clustering (marked versus unmarked)

The difference in excitation parameters *α* and *β* were negligible across the marked and unmarked specifications (Cohen’s *d* < 0.2 in both cohorts; Figure 4), indicating that the timescale and strength of self-excitation are insensitive to intensity marks. The derived branching ratio *n* = *α/β* shifted somewhat more (hip *d* = 0.693; wrist *d* = 0.843), but this may reflect re-balancing between *µ* and the excitation term under mark normalization rather than a structural change; marks also widened the cross-participant distributions of *µ* and *n*, consistent with mark-shape heterogeneity adding a between-individual source of variation. Incorporating marks improved GOF only slightly (KS *d* = 0.32). The empirical equivalence of (*α, β*) supports describing temporal clustering by event timing alone.

**Figure 4:**
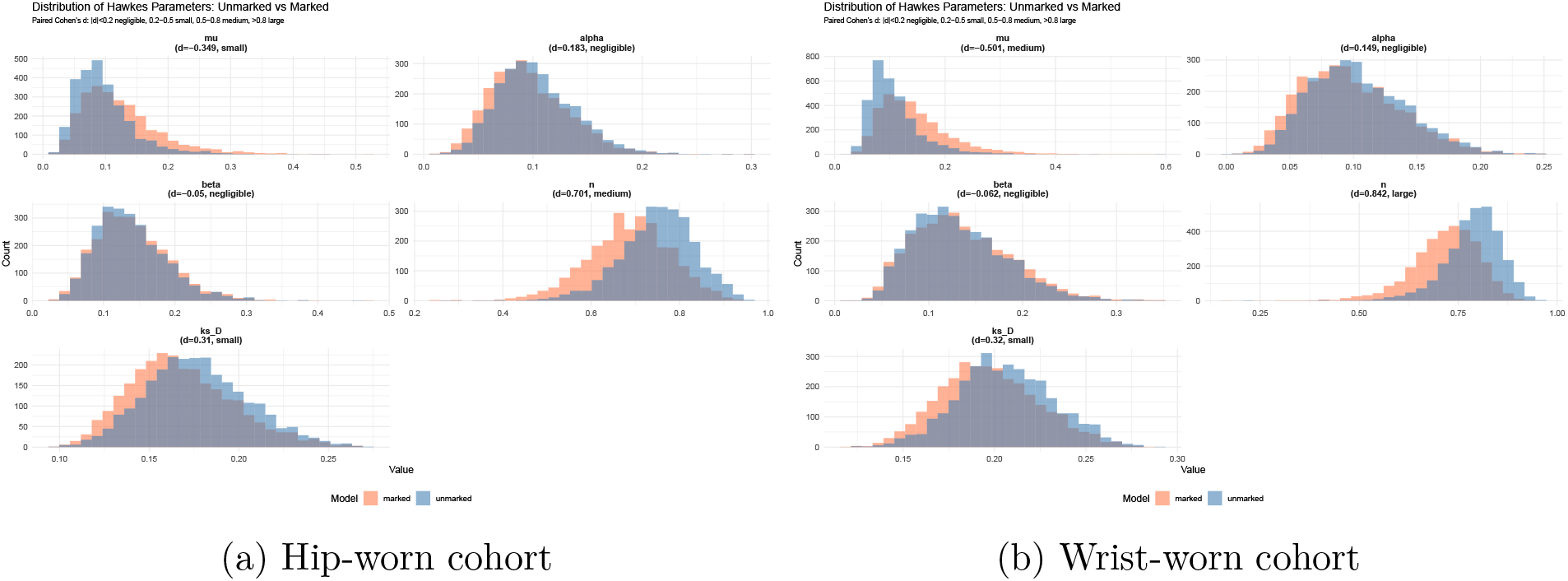
Distribution of Hawkes parameters from maximum likelihood estimation (MLE): unmarked (blue) versus marked (orange) models. Paired Cohen’s *d* effect sizes are shown. The kernel parameters *α* (excitation strength) and *β* (decay rate) showed negligible differences (|*d*| < 0.2).

### 5.4 Agreement between dispersion-based and MLE estimators

Across participants (Figure 5), the marked-model MLE branching ratio averaged 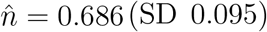 for hip and 0.718 (SD 0.084) for wrist—roughly 70% of active epochs arising from endogenous persistence rather than exogenous initiation. The mean event rates was 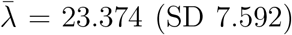 for hip-worn and 30.143 (SD 7.255) events per hour for wrist-worn data.

**Figure 5:**
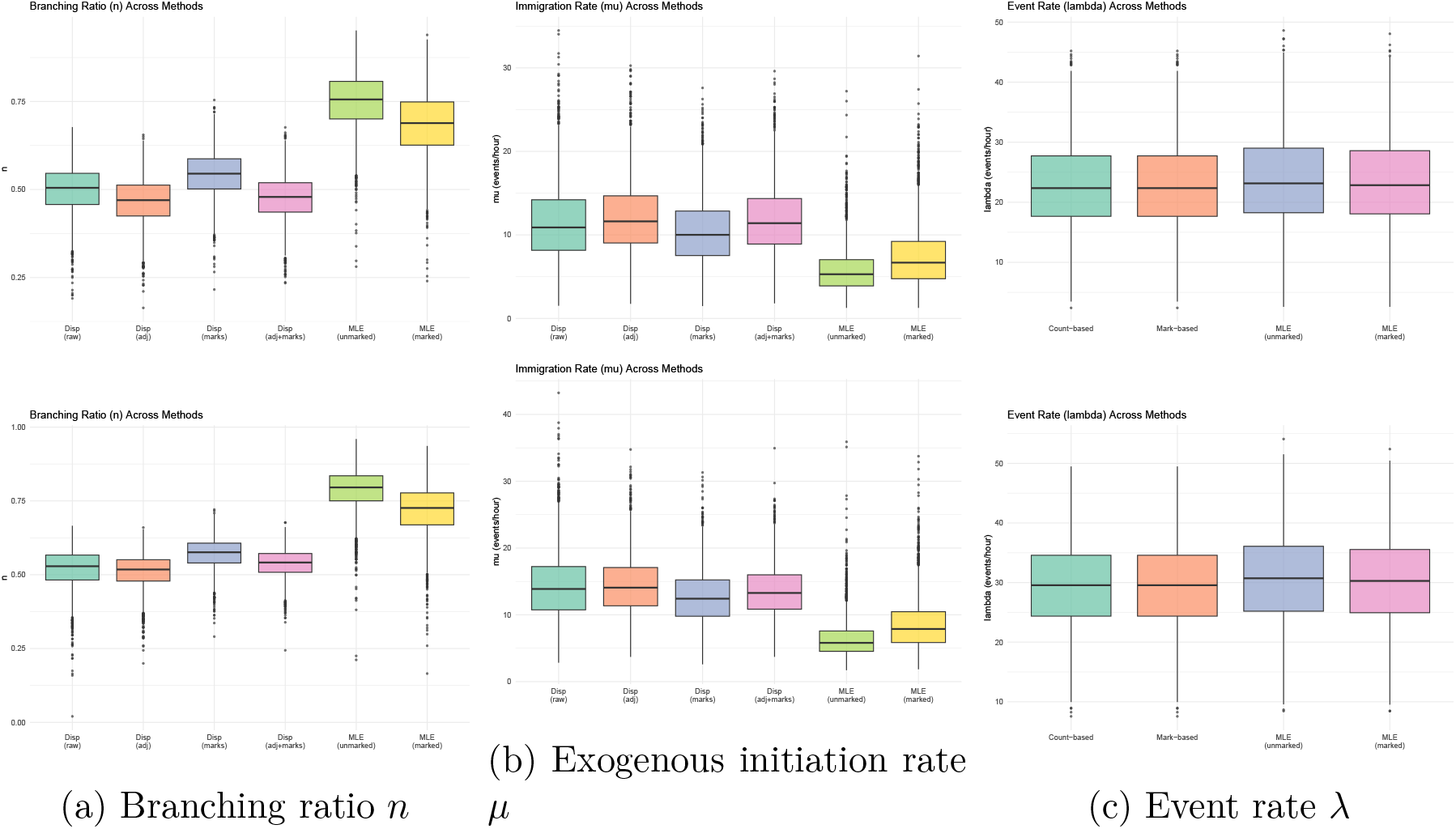
Distribution of MLE and dispersion-based parameter estimates across participants. Top row: hip-worn cohort; bottom row: wrist-worn cohort.

The correlations between dispersion-based parameters, MLE estimates, and traditional PA metrics are in Supplementary Figure 20. The event-count-based dispersion index correlated strongly with the MLE branching ratios (*r* ≈ 0.74 in both cohorts; Figure 6), which validated the moment estimator as a far cheaper alternative to likelihood estimation. The intensity-marked dispersion index agreed less well (*r* = 0.48–0.65), indicating that high intensity variance injects noise that partly obscures the clustering signal; together with the marked-versus-unmarked equivalence of *α* and *β*, this favours the event-count-based over the intensity-mark-weighted estimator. The moderate negative correlation between *n* and *µ* matched the stationarity constraint 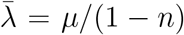, reflecting a trade-off between exogenous initiation and endogenous persistence at a given activity rate. Finally, the branching ratio correlated only weakly with conventional PA metrics ( |*r*| ≤ 0.50 for total activity volume, Peak30, ASTP, SATP, wear time, MVPA, and sedentary time), confirming that it captures information largely distinct from volume; the event rate *λ*, by contrast, correlated moderately-to-strongly with several volume metrics and with Peak30. We therefore selected Peak30 as the single volume/intensity covariate in the survival models, summarizing the most active part of the day while avoiding the multicollinearity of including several PA metrics.

**Figure 6:**
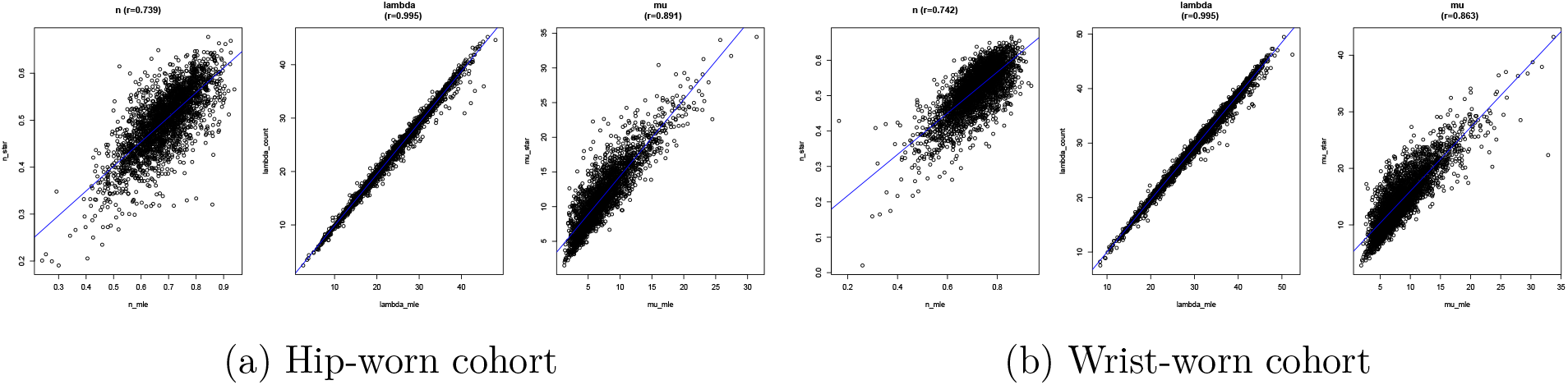
Scatter plots showing correlations between dispersion-based and MLE estimates of the branching ratio *n*, baseline rate *µ*, and event rate *λ*.

### 5.5 Temporal clustering and all-cause mortality

Having established a scalable, inexpensive dispersion-based estimator of the branching ratio *n*, we asked whether it carries prognostic information, relating *n* and *λ* to all-cause mortality across all six estimators (Tables 3 and 4).

**Table 3:** PA temporal dynamics and all-cause mortality in the hip-worn cohort (2003-2006). Hazard ratio (HR) with 95% confidence interval (CI) per standard deviation (SD).

| Method | $n$ (M1) | $\lambda$ (M1) | $n$ (M2) | $\lambda$ (M2) | Peak30 (M2) |
| --- | --- | --- | --- | --- | --- |
| Hawkes MLE (unmarked) | 0.750 (0.664–0.847)*** | 0.760 (0.652–0.887)*** | 0.849 (0.739–0.976)* | 0.944 (0.803–1.111) | 0.622 (0.528–0.732)*** |
| Hawkes MLE (marked) | 0.829 (0.720–0.954)** | 0.686 (0.589–0.800)*** | 0.833 (0.722–0.961)* | 0.923 (0.792–1.076) | 0.592 (0.506–0.692)*** |
| Dispersion (event-count-based) | 0.702 (0.634–0.777)*** | 0.683 (0.593–0.786)*** | 0.819 (0.725–0.926)** | 0.881 (0.765–1.015) | 0.633 (0.537–0.747)*** |
| Dispersion (TOD-adj event-count) | 0.779 (0.711–0.854)*** | 0.730 (0.628–0.848)*** | 0.882 (0.803–0.968)** | 0.923 (0.794–1.073) | 0.617 (0.525–0.727)*** |
| Dispersion (intensity-mark-weighted) | 0.700 (0.635–0.772)*** | 0.742 (0.644–0.855)*** | 0.844 (0.746–0.955)** | 0.896 (0.775–1.036) | 0.659 (0.555–0.782)*** |
| Dispersion (TOD-adj intensity-mark) | 0.743 (0.674–0.818)*** | 0.763 (0.658–0.884)*** | 0.877 (0.791–0.973)* | 0.924 (0.794–1.074) | 0.635 (0.539–0.750)*** |

Model 1 (M1): age, sex, race, and wear time (WT). Model 2 (M2): M1 plus Peak30, education, BMI, smoking, drinking, and comorbidities.

**Table 4:** PA temporal dynamics and all-cause mortality in the wrist-worn cohort (2011-2014). Hazard ratio (HR) with 95% confidence interval (CI) per standard deviation (SD).

| Method | $n$ (M1) | $\lambda$ (M1) | $n$ (M2) | $\lambda$ (M2) | Peak30 (M2) |
| --- | --- | --- | --- | --- | --- |
| Hawkes MLE (unmarked) | 0.928 (0.776–1.108) | 0.696 (0.588–0.823)*** | 1.052 (0.917–1.206) | 1.007 (0.809–1.252) | 0.492 (0.362–0.670)*** |
| Hawkes MLE (marked) | 1.063 (0.897–1.259) | 0.687 (0.576–0.818)*** | 1.099 (0.951–1.271) | 1.006 (0.814–1.243) | 0.504 (0.365–0.695)*** |
| Dispersion (event-count-based) | 0.847 (0.706–1.018) | 0.650 (0.525–0.805)*** | 1.101 (0.946–1.283) | 1.073 (0.838–1.374) | 0.477 (0.348–0.655)*** |
| Dispersion (TOD-adj event-count) | 1.127 (0.963–1.319) | 0.687 (0.579–0.815)*** | 1.309 (1.127–1.521)*** | 1.043 (0.849–1.282) | 0.449 (0.327–0.617)*** |
| Dispersion (intensity-mark-weighted) | 0.768 (0.654–0.902)** | 0.649 (0.536–0.786)*** | 1.046 (0.902–1.213) | 1.046 (0.810–1.350) | 0.479 (0.341–0.671)*** |
| Dispersion (TOD-adj intensity-mark) | 1.017 (0.875–1.181) | 0.688 (0.585–0.810)*** | 1.218 (1.068–1.388)** | 1.028 (0.833–1.270) | 0.443 (0.314–0.626)*** |

Model 1 (M1): age, sex, race, and wear time (WT). Model 2 (M2): M1 plus Peak30, education, BMI, smoking, drinking, and comorbidities.

In the hip-worn cohort, higher PA temporal clustering was consistently associated with lower mortality. In Model 1, hazard ratios (HRs) ranged from 0.700 to 0.829 per SD (all *p* < 0.01), corresponding to 17–30% lower risk per SD, and after adjustment for Peak30 in Model 2 the association remained (HR 0.819–0.882, *p* < 0.05). This indicates that temporal clustering predicts mortality with partial independence from frequency and intensity-driven volume. The event rate *λ* was protective in Model 1 (HR 0.683–0.763, all *p* < 0.001) but attenuated to the null in Model 2, suggesting that frequency is largely accounted for by intensity-driven volume. Marked and unmarked specifications gave similar HRs, consistent with the finding that PA intensity does not alter the temporal clustering patterns.

In the wrist-worn cohort, the estimator behaved identically at every stage upstream of the mortality analysis—the validation against MLE, the recovered *n* ≈ 0.75, the GOF, and the marked-versus-unmarked similarity—yet the direction of the mortality association reversed: after Peak30 adjustment all six estimators gave HRs above one (Table 4), reaching significance under the two TOD-adjusted specifications. As in the hip cohort, *λ* was protective in Model 1 but null in Model 2, and the marked and unmarked models agreed. Peak30 was strongly protective in both cohorts (HR 0.443–0.659, all *p* < 0.001).

The dispersion index is closely related to the coefficient of variation (CoV) through *D* = CoV^2^ · *µ*—both quantify variability relative to mean activity. A supplementary analysis of nocturnal activity (restricted to the wrist cohort, since the hip protocol removed the device at night) found that lower CoV of 1–5 a.m. activity—more consolidated nocturnal behavior— was associated with lower mortality (HR 0.645, *p* = 0.029), whereas mean nocturnal volume was not (Supplementary Material II, Table 2).

## 6 Discussion

Using a scalable, kernel-independent estimator of the Hawkes branching ratio—an alternative to maximum likelihood estimates—we characterized temporal clustering in human physical activity (PA). Three findings emerged. First, TOD-adjusted dispersion indices exceeded 1 for nearly all participants, confirming persistent clustering consistent with self-excitation beyond TOD effects. Second, marked and unmarked models yielded equivalent excitation and decay parameters with comparable goodness of fit, indicating that PA intensity does not modulate the clustering structure. Third, the branching ratio was associated with all-cause mortality independently of activity frequency and intensity-driven volume, suggesting that temporal clustering carries health information beyond total volume.

### Methodological context

Human activities are well documented to follow non-Poisson statistics: bursts of rapidly occurring events separated by long quiescent periods, across online and offline human activities such as communication, study and work [34, 49, 50, 51, 52]. The burstiness framework offers features particularly relevant for PA research: temporal clustering not fully captured by aggregate measures, within-bout dynamics driven by epoch-level self-triggering, and memory effects indicating the extent to which one active epoch influences the timing of subsequent epochs. Our finding that PA onsets are overdispersed with positive branching ratios confirms that accelerometry-derived active epochs follow the same bursty pattern, and the branching ratio quantifies it directly—the proportion of active epochs endogenously triggered by preceding activity rather than by exogenous cues.

This mechanistic perspective distinguishes the branching ratio from existing approaches that go beyond volume and bout summaries. Metrics of absolute timing (e.g., the cosinor acrophase [53]) capture *when* activity peaks, and regularity metrics such as sample entropy [54] and CoV summarize *how variable* activity is; neither decomposes observed temporal variability into exogenous and endogenous components, as the branching ratio and dispersion-based index do. The term “dispersion” warrants clarification: Klamm *et al*. [55] use “movement dispersion” for activity spread evenly across the day, whereas our point-process usage takes *D* > 1 to mean temporal clustering (overdispersion)—shared terminology, fundamentally different aspects of PA patterning.

The moment-based dispersion approach requires only mean and variance calculations, making it computationally straightforward for large-scale accelerometry datasets, and the strong correlation (*r* ≈ 0.74) between event-count-based dispersion estimates and maximum likelihood estimates validates this simpler approach. The TOD-adjusted dispersion specifications showed weaker correlations with MLE-based estimates. Under approximate stationarity, TOD detrending removes deterministic exogenous variation, reducing agreement with the MLE branching ratio but isolating the endogenous clustering component more directly relevant to health outcomes.

### Clinical implications

Device-measured PA volume shows a well-established dose–response association with all-cause mortality [2, 1, 56]. Beyond volume, temporal structure such as PA fragmentation and sleep regularity carries independent prognostic information: [57, 58]. Our association between temporal clustering (*n*) and mortality, after adjustment for event rate (*λ*) and Peak30, fits this pattern.

The equivalence of marked and unmarked models suggests the benefit of temporal clustering comes from maintaining the active state rather than from intensity, consistent with evidence that activity of any intensity—and a consistent day-to-day routine—lowers mortality risk [44, 2]. Interventions may therefore benefit from supporting sustained activity once initiated, regardless of intensity level. The decomposition into exogenous initiation (*µ*^∗^) and endogenous persistence (*n*^∗^) further points to individualized intervention targets: low *µ*^∗^ with adequate *n*^∗^ may call for environmental prompts and scheduling, while adequate *µ*^∗^ with low *n*^∗^ may call for pacing and social support. Rather than relying on generic recommendations to increase MVPA, public health approaches could be tailored according to whether individuals experience greater difficulty initiating activity, sustaining activity, or both.

Associations differed by sensor placement: the hip-worn branching ratio was consistently protective, whereas wrist-derived estimates were inconsistent in direction—echoing Zhou *et al*. [44], who found that indicators from the same MIMS threshold can yield divergent mortality associations depending on how temporal structure is summarized. This between-placement discrepancy warrants caution, because the hip and wrist data come from different cohorts differing in demographics, sample size, study design, and follow-up length, so it may reflect cohort differences as much as what each sensor captures. Mechanistically, hip accelerometry mainly records voluntary ambulation, a more specific measure of purposeful movement, whereas the wrist also captures incidental upper-limb motion that may add variability. In addition, differing sedentary thresholds across calibration frameworks could further contribute.

### Limitations

Several limitations remain. First, the dispersion estimator was computed per day and averaged, whereas the MLE shared parameters across days, so between-day variation in baseline rate was not modeled; given evidence that concentrating activity into one or two weekly sessions confers comparable benefit [44, 59, 60], day-specific baselines are worth exploring. Second, we assumed intensity-marks were conditionally independent of the process, leaving nonlinear or time-dependent mark functions unexplored. Third, although most daily series passed Ljung–Box, a minority did not across all seven days, and Q–Q plots showed some departures from Exp(1)—likely reflecting heavy tails, intra-day regime changes, or extreme bursts that a single exponential kernel cannot absorb—motivating richer (power-law [61] or multi-exponential) kernels in future work. Finally, the TOD adjustment is an empirical detrending step rather than a result derived within the stationary theory; we report the adjusted estimates only as a companion to the unadjusted ones, and a formal treatment of estimation under non-stationarity remains open.

## 7 Conclusions

We extended the dispersion-based branching ratio estimatora parsimonious, kernel-independent method for quantifying temporal clusteringto marked Hawkes processes, enabling its application to accelerometry data in which activity intensity varies substantially, and demonstrated that activity intensity does not alter the underlying temporal excitation structure. From a clinical perspective, these findings suggest that the benefits of temporal clustering are more closely related to persistence of the active state than to maintenance of high activity intensity. This interpretation provides actionable targets for improving both activity initiation and sustainability, potentially extending the health benefits obtained from overall activity accumulation.

## Data Availability

This study used publicly available NHANES 2003--2006 and 2011--2014 data (\url{https://www.cdc.gov/nchs/nhanes/}). Analysis code is available at \url{https://github.com/QuQ-ToT-Orz/HUO_Project2}.

## Declarations

### Ethics approval and consent to participate

This study used publicly available, de-identified NHANES data; ethics approval was not required for secondary analysis of anonymized public data.

### Availability of data and materials

This study used publicly available NHANES 2003– 2006 and 2011–2014 data (https://www.cdc.gov/nchs/nhanes/). Analysis code is available at https://github.com/QuQ-ToT-Orz/HUO_Project2. NHANES 2003–2006 data were processed using the R package `rnhanesdata;` analogous functions and a pipeline for 2011– 2014 were developed in the R package `HUO` (Hawkes-based Unfolding of Observed human behavior), which implements shared-parameter Hawkes estimation with branching-ratio penalization, drawing on the `stelfi` framework. The `HUO` package and full reproduction pipeline are available at the same repository.

### Authors’ contributions

X.Z. conceived and designed the study, developed the methodology, performed the statistical analyses, and drafted the manuscript. I.D. and M.C. critically reviewed the manuscript and provided substantial feedback. All authors read and approved the final manuscript.

## Acknowledgements

The authors thank Dr. Deborah Sulem (Università della Svizzera Italiana) for valuable discussions on Hawkes process modeling and framing, and Dr. Wessel van Wieringen for early discussions on parameter regularization.

## Abbreviations

ASTP, active-to-sedentary transition probability; BMI, body mass index; CI, confidence interval; CoV, coefficient of variation; TOD, time-of-day; GOF, goodness of fit; HDL, high-density lipoprotein; HR, hazard ratio; KS, Kolmogorov–Smirnov; MIMS, Monitor-Independent Movement Summary; MLE, maximum likelihood estimation; MVPA, moderate-to-vigorous physical activity; NHANES, National Health and Nutrition Examination Survey; PA, physical activity; Peak30, average of the 30 highest 1-minute activity values per day; SATP, sedentary-to-active transition probability; SD, standard deviation; ST, sedentary time; TAC, total activity counts; WT, wear time.

## References

[1] Edvard H Sagelv et al. “Device-measured physical activity, sedentary time, and risk of all-cause mortality: an individual participant data analysis of four prospective cohort studies”. en. In: British Journal of Sports Medicine 57.22 (Nov. 2023), pp. 1457–1463. ISSN: 0306-3674, 1473-0480. DOI: 10.1136/bjsports-2022-106568.

[2] Ulf Ekelund et al. “Dose-response associations between accelerometry measured physical activity and sedentary time and all cause mortality: systematic review and harmonised meta-analysis”. en. In: BMJ (Aug. 2019), p. 4570. ISSN: 0959-8138, 1756-1833. DOI: 10.1136/bmj.l4570.

[3] Fiona C Bull et al. “World Health Organization 2020 guidelines on physical activity and sedentary behaviour”. en. In: British Journal of Sports Medicine 54.24 (Dec. 2020), pp. 1451–1462. ISSN: 0306-3674, 1473-0480. DOI: 10.1136/bjsports-2020-102955.

[4] Tessa Strain et al. “Wearable-device-measured physical activity and future health risk”. en. In: Nature Medicine 26.9 (Sept. 2020), pp. 1385–1391. ISSN: 1078-8956, 1546-170X. DOI: 10.1038/s41591-020-1012-3.

[5] Ya-Ting Liang, Charlotte Wang, and Chuhsing Kate Hsiao. “Data Analytics in Physical Activity Studies With Accelerometers: Scoping Review”. en. In: Journal of Medical Internet Research 26 (Sept. 2024), e59497. ISSN: 1438-8871. DOI: 10.2196/59497.

[6] Qiang Wang et al. “Does Physical Activity Intensity Matter? Longitudinal Evidence From a 5-Wave National Chinese Cohort on Chronic Disease Prevalence”. In: Journal of Physical Activity and Health 22.12 (Dec. 2025), pp. 1602–1610. ISSN: 1543-3080, 1543-5474. DOI: 10.1123/jpah.2025-0284.

[7] Elsa Kobeissi et al. “A Systematic Review and Taxonomy of Metrics and Methods Used to Describe Temporal Movement Behavior Patterns Measured Using Wearable Devices”. In: Journal for the Measurement of Physical Behaviour 9.1 (Jan. 2026), jmpb.2024–0035. ISSN: 2575-6605, 2575-6613. DOI: 10.1123/jmpb.2024-0035.

[8] Catrine Tudor-Locke et al. “How fast is fast enough? Walking cadence (steps/min) as a practical estimate of intensity in adults: a narrative review”. en. In: British Journal of Sports Medicine 52.12 (June 2018), pp. 776–788. ISSN: 0306-3674, 1473-0480. DOI: 10.1136/bjsports-2017-097628.

[9] Emmanuel Stamatakis et al. “Association of wearable device-measured vigorous intermittent lifestyle physical activity with mortality”. en. In: Nature Medicine 28.12 (Dec. 2022), pp. 2521–2529. ISSN: 1078-8956, 1546-170X. DOI: 10.1038/s41591-022-02100-x.

[10] Ulf Ekelund et al. “Joint associations of accelerometer-measured physical activity and sedentary time with all-cause mortality: a harmonised meta-analysis in more than 44 000 middle-aged and older individuals”. en. In: British Journal of Sports Medicine 54.24 (Dec. 2020), pp. 1499–1506. ISSN: 0306-3674, 1473-0480. DOI: 10.1136/bjsports-2020-103270.

[11] Sarah Kozey Keadle et al. “An Evaluation of Accelerometer-derived Metrics to Assess Daily Behavioral Patterns”. In: Medicine & Science in Sports & Exercise 49.1 (Jan. 2017), pp. 54–63. ISSN: 0195-9131. DOI: 10.1249/MSS.0000000000001073.

[12] W. Witting et al. “Alterations in the circadian rest-activity rhythm in aging and Alzheimer’s disease”. en. In: Biological Psychiatry 27.6 (Mar. 1990), pp. 563–572. ISSN: 00063223. DOI: 10.1016/0006-3223(90)90523-5.

[13] Eus J. W. Van Someren et al. “Bright Light Therapy: Improved Sensitivity to Its Effects on Rest-Activity Rhythms in Alzheimer Patients by Application of Nonparametric Methods”. en. In: Chronobiology International 16.4 (Jan. 1999), pp. 505–518. ISSN: 0742-0528, 1525-6073. DOI: 10.3109/07420529908998724.

[14] Els Weinans et al. “Signals of complexity and fragmentation in accelerometer data”. In: PLOS One 20.7 (July 2025). Ed. by Sandip V. George, e0326522. ISSN: 1932-6203. DOI: 10.1371/journal.pone.0326522.

[15] S.F.M. Chastin and M.H. Granat. “Methods for objective measure, quantification and analysis of sedentary behaviour and inactivity”. en. In: Gait & Posture 31.1 (Jan. 2010), pp. 82–86. ISSN: 09666362. DOI: 10.1016/j.gaitpost.2009.09.002.

[16] Anisoara Paraschiv-Ionescu et al. “Barcoding Human Physical Activity to Assess Chronic Pain Conditions”. en. In: PLoS ONE 7.2 (Feb. 2012). Ed. by Ken R. Duffy, e32239. ISSN: 1932-6203. DOI: 10.1371/journal.pone.0032239.

[17] Junrui Di et al. Patterns of sedentary and active time accumulation are associated with mortality in US adults: The NHANES study. Aug. 2017. DOI: 10.1101/182337.

[18] Ian Meneghel Danilevicz et al. “Measures of fragmentation of rest activity patterns: mathematical properties and interpretability based on accelerometer real life data”. en. In: BMC Medical Research Methodology 24.1 (June 2024), p. 132. ISSN: 1471-2288. DOI: 10.1186/s12874-024-02255-w.

[19] Simone T Boerema et al. “Pattern measures of sedentary behaviour in adults: A literature review”. en. In: DIGITAL HEALTH 6 (Jan. 2020), p. 2055207620905418. ISSN: 2055-2076, 2055-2076. DOI: 10.1177/2055207620905418.

[20] Keerati Suibkitwanchai et al. “Nonparametric time series summary statistics for high-frequency accelerometry data from individuals with advanced dementia”. en. In: PLOS ONE 15.9 (Sept. 2020). Ed. by j E. Trinidad Segovia, e0239368. ISSN: 1932-6203. DOI: 10.1371/journal.pone.0239368.

[21] Alan G. Hawkes. “Spectra of some self-exciting and mutually exciting point processes”. en. In: Biometrika 58.1 (1971), pp. 83–90. ISSN: 0006-3444, 1464-3510. DOI: 10.1093/biomet/58.1.83.

[22] Waltenegus Dargie. “Analysis of Time and Frequency Domain Features of Accelerometer Measurements”. In: 2009 Proceedings of 18th International Conference on Computer Communications and Networks. IEEE, Aug. 2009, pp. 1–6. DOI: 10.1109/ICCCN.2009.5235366.

[23] Daryl J. Daley and D. Vere-Jones. An introduction to the theory of point processes. 2nd ed. New York: Springer, 2003. isbn: 978-0-387-95541-4 978-0-387-21337-8 978-0-387-49835-5.

[24] Yosihiko Ogata. “Statistical Models for Earthquake Occurrences and Residual Analysis for Point Processes”. en. In: Journal of the American Statistical Association 83.401 (Mar. 1988), pp. 9–27. ISSN: 0162-1459, 1537-274X. DOI: 10.1080/01621459.1988.10478560.

[25] Patrick J. Laub et al. “Hawkes Models and Their Applications”. en. In: Annual Review of Statistics and Its Application 12.1 (Mar. 2025), pp. 233–258. ISSN: 2326-8298, 2326-831X. DOI: 10.1146/annurev-statistics-112723-034304.

[26] Rafael Lima. “Hawkes Processes Modeling, Inference, and Control: An Overview”. en. In: SIAM Review 65.2 (May 2023), pp. 331–374. ISSN: 0036-1445, 1095-7200. DOI: 10.1137/21M1396927.

[27] Wen-Hao Chiang, Xueying Liu, and George Mohler. “Hawkes process modeling of COVID-19 with mobility leading indicators and spatial covariates”. en. In: International Journal of Forecasting 38.2 (Apr. 2022), pp. 505–520. ISSN: 01692070. DOI: 10.1016/j.ijforecast.2021.07.001.

[28] Alba Bernabeu, Jiancang Zhuang, and Jorge Mateu. “Spatio-Temporal Hawkes Point Processes: A Review”. en. In: Journal of Agricultural, Biological and Environmental Statistics 30.1 (Mar. 2025), pp. 89–119. ISSN: 1085-7117, 1537-2693. DOI: 10.1007/s13253-024-00653-7.

[29] Anisoara Paraschiv-Ionescu, Eric Buchser, and Kamiar Aminian. “Unraveling dynamics of human physical activity patterns in chronic pain conditions”. en. In: Scientific Reports 3.1 (June 2013), p. 2019. ISSN: 2045-2322. DOI: 10.1038/srep02019.

[30] Emmanuel Bacry, Iacopo Mastromatteo, and Jean-François Muzy. Hawkes processes in finance. en. arXiv:1502.04592 [q-fin]. May 2015. DOI: 10.48550/arXiv.1502.04592.

[31] V. Filimonov and D. Sornette. “Apparent criticality and calibration issues in the Hawkes self-excited point process model: application to high-frequency financial data”. en. In: Quantitative Finance 15.8 (Aug. 2015), pp. 1293–1314. ISSN: 1469-7688, 1469-7696. DOI: 10.1080/14697688.2015.1032544.

[32] Stephen J. Hardiman, Nicolas Bercot, and Jean-Philippe Bouchaud. “Critical reflexivity in financial markets: a Hawkes process analysis”. en. In: The European Physical Journal B 86.10 (Oct. 2013), p. 442. ISSN: 1434-6028, 1434-6036. DOI: 10.1140/epjb/e2013-40107-3.

[33] Mehdi Lallouache and Damien Challet. “The limits of statistical significance of Hawkes processes fitted to financial data”. en. In: Quantitative Finance 16.1 (Jan. 2016), pp. 1–11. ISSN: 1469-7688, 1469-7696. DOI: 10.1080/14697688.2015.1068442.

[34] Albert-László Barabási. “The origin of bursts and heavy tails in human dynamics”. In: Nature 435.7039 (May 2005). arXiv: cond-mat/0505371, pp. 207–211. ISSN: 0028-0836, 1476-4687. DOI: 10.1038/nature03459.

[35] Stephen J. Hardiman and Jean-Philippe Bouchaud. “Branching-ratio approximation for the self-exciting Hawkes process”. en. In: Physical Review E 90.6 (Dec. 2014), p. 062807. ISSN: 1539-3755, 1550-2376. DOI: 10.1103/PhysRevE.90.062807.

[36] Andrew Leroux et al. “Organizing and Analyzing the Activity Data in NHANES”. In: Statistics in Biosciences 11.2 (July 2019), pp. 262–287. ISSN: 1867-1764, 1867-1772. DOI: 10.1007/s12561-018-09229-9.

[37] Dinesh John et al. “An Open-Source Monitor-Independent Movement Summary for Accelerometer Data Processing”. In: Journal for the Measurement of Physical Behaviour 2.4 (Dec. 2019), pp. 268–281. ISSN: 2575-6605, 2575-6613. DOI: 10.1123/jmpb.2018-0068.

[38] T. Ozaki. “Maximum likelihood estimation of Hawkes’ self-exciting point processes”. In: Annals of the Institute of Statistical Mathematics 31.1 (Dec. 1979), pp. 145–155. ISSN: 0020-3157, 1572-9052. DOI: 10.1007/BF02480272.

[39] Kamil Rajdl, Petr Lansky, and Lubomir Kostal. “Fano Factor: A Potentially Useful Information”. In: Frontiers in Computational Neuroscience 14 (Nov. 2020), p. 569049. ISSN: 1662-5188. DOI: 10.3389/fncom.2020.569049.

[40] Alan G. Hawkes and David Oakes. “A cluster process representation of a self-exciting process”. en. In: Journal of Applied Probability 11.3 (Sept. 1974), pp. 493–503. ISSN: 0021-9002, 1475-6072. DOI: 10.2307/3212693.

[41] Fraser Daly and Seva Shneer. The Borel Distribution: Approximation and Concentration. en. arXiv:1912.03219 [math.PR]. July 2021. DOI: 10.48550/arXiv.1912.03219.

[42] Richard P. Troiano et al. “Physical Activity in the United States Measured by Accelerometer”. In: Medicine & Science in Sports & Exercise 40.1 (Jan. 2008), pp. 181–188. ISSN: 0195-9131. DOI: 10.1249/mss.0b013e31815a51b3.

[43] Marta Karas et al. Comparison of Accelerometry-based Measures of Physical Activity. Mar. 2022. DOI: 10.1101/2022.03.16.22272518.

[44] Xinkai Zhou et al. Generalized Multilevel Functional Principal Component Analysis with Application to NHANES Active Inactive Patterns. en. arXiv:2311.14054 [stat]. Mar. 2025. DOI: 10.48550/arXiv.2311.14054.

[45] Annemarie Koster et al. “Comparison of Sedentary Estimates between activPAL and Hip- and Wrist-Worn ActiGraph”. In: Medicine & Science in Sports & Exercise 48.8 (Aug. 2016), pp. 1514–1522. ISSN: 0195-9131. DOI: 10.1249/MSS.0000000000000924.

[46] I. Rubin. “Regular point processes and their detection”. In: IEEE Transactions on Information Theory 18.5 (Sept. 1972), pp. 547–557. ISSN: 0018-9448, 1557-9654. DOI: 10.1109/TIT.1972.1054897.

[47] Emery N. Brown et al. “The Time-Rescaling Theorem and Its Application to Neural Spike Train Data Analysis”. en. In: Neural Computation 14.2 (Feb. 2002), pp. 325–346. ISSN: 0899-7667, 1530-888X. DOI: 10.1162/08997660252741149.

[48] Kyungsub Lee. emhawkes: Exponential Multivariate Hawkes Model. R package version 0.9.8. 2025.

[49] Alexei Vázquez et al. “Modeling bursts and heavy tails in human dynamics”. en. In: Physical Review E 73.3 (Mar. 2006), p. 036127. ISSN: 1539-3755, 1550-2376. DOI: 10. 1103/PhysRevE.73.036127.

[50] Márton Karsai, Hang-Hyun Jo, and Kimmo Kaski. Bursty Human Dynamics. en. SpringerBriefs in Complexity. Cham: Springer International Publishing, 2018. isbn: 978-3-319-68538-0 978-3-319-68540-3. DOI: 10.1007/978-3-319-68540-3.

[51] Zhi-Dan Zhao et al. “Empirical Analysis on the Human Dynamics of a Large-Scale Short Message Communication System”. In: Chinese Physics Letters 28.6 (June 2011), p. 068901. ISSN: 0256-307X, 1741-3540. DOI: 10.1088/0256-307X/28/6/068901.

[52] K.-I. Goh and A.-L. Barabási. “Burstiness and memory in complex systems”. In: EPL (Europhysics Letters) 81.4 (Feb. 2008), p. 48002. ISSN: 0295-5075, 1286-4854. DOI: 10.1209/0295-5075/81/48002.

[53] Germaine Cornelissen. “Cosinor-based rhythmometry”. en. In: Theoretical Biology and Medical Modelling 11.1 (Dec. 2014), p. 16. ISSN: 1742-4682. DOI: 10.1186/1742-4682-11-16.

[54] Joshua S. Richman and J. Randall Moorman. “Physiological time-series analysis using approximate entropy and sample entropy”. en. In: American Journal of Physiology-Heart and Circulatory Physiology 278.6 (June 2000), H2039–H2049. ISSN: 0363-6135, 1522-1539. DOI: 10.1152/ajpheart.2000.278.6.H2039.

[55] Melissa M. Klamm et al. “Dispersion of daily physical activity behaviors in schoolage children: A novel approach to measure patterns of physical activity”. en. In: Journal for Specialists in Pediatric Nursing 27.2 (Apr. 2022), e12364. ISSN: 1539-0136, 1744-6155. DOI: 10.1111/jspn.12364.

[56] Ulf Ekelund et al. “Deaths potentially averted by small changes in physical activity and sedentary time: an individual participant data meta-analysis of prospective cohort studies”. en. In: The Lancet 407.10526 (Jan. 2026), pp. 339–349. ISSN: 01406736. DOI: 10.1016/S0140-6736(25)02219-6.

[57] Mathilde Chen et al. “Identification of physical activity and sedentary behaviour dimensions that predict mortality risk in older adults: development of a machine learning model in the Whitehall II accelerometer sub-study and external validation in the CoLaus study”. en. In: eClinicalMedicine 55 (Jan. 2023), p. 101773. ISSN: 25895370. DOI: 10.1016/j.eclinm.2022.101773.

[58] Daniel P Windred et al. “Sleep regularity is a stronger predictor of mortality risk than sleep duration: A prospective cohort study”. en. In: SLEEP 47.1 (Jan. 2024), zsad253. ISSN: 0161-8105, 1550-9109. DOI: 10.1093/sleep/zsad253.

[59] Gary ODonovan et al. “Association of Weekend Warrior and Other Leisure Time Physical Activity Patterns With Risks for All-Cause, Cardiovascular Disease, and Cancer Mortality”. en. In: JAMA Internal Medicine 177.3 (Mar. 2017), p. 335. ISSN: 2168-6106. DOI: 10.1001/jamainternmed.2016.8014.

[60] Mauricio Dos Santos et al. “Association of the Weekend Warrior and Other Leisure-time Physical Activity Patterns With All-Cause and Cause-Specific Mortality: A Nationwide Cohort Study”. en. In: JAMA Internal Medicine 182.8 (Aug. 2022), p. 840. ISSN: 2168-6106. DOI: 10.1001/jamainternmed.2022.2488.

[61] Aaron Clauset, Cosma Rohilla Shalizi, and M. E. J. Newman. “Power-Law Distributions in Empirical Data”. en. In: SIAM Review 51.4 (Nov. 2009), pp. 661–703. ISSN: 0036-1445, 1095-7200. DOI: 10.1137/070710111.

