## Supplementary material for "Scalable estimation of temporal clustering in accelerometry: a kernel-independent dispersion index grounded in the Hawkes process": S1

### Supplementary Material I:
### Derivation of the Marked Hawkes Process Dispersion Index Estimator

Xin Zheng*[1,2], Ian Meneghel Danilevicz[3], and Maichina Paw[1,2]

[1]Amsterdam UMC location Vrije Universiteit Amsterdam, Public and Occupational Health, De Boelelaan 1117, Amsterdam, the Netherlands
[2]Amsterdam Public Health, Health Behaviors and Chronic Diseases and Methodology, Amsterdam, the Netherlands
[3]Université Paris Cité, Inserm U1153, Centre for Research in Epidemiology and Statistics (CRESS), Epidemiology of Ageing and Neurodegenerative Diseases (EpiAgeing), Paris, France

#### 1 Introduction

This supplementary material provides a complete mathematical derivation of the dispersion-based branching ratio estimator for marked Hawkes processes. We extend the kernel-independent approach of Hardiman & Bouchaud [1] by explicitly accounting for the variance of event marks (physical activity intensities). The derivation shows the relationship $D_{\text{marked}} = \mathcal{Q} \cdot 1/(1-n)^2$, where the mark-moment ratio $\mathcal{Q} = E[M^2]/E[M]$ emerges naturally from the cluster representation of marked point processes.

#### 2 Model Setup and Assumptions

##### 2.1 The Marked Hawkes Process

**Assumption 1** (Stationary marked Hawkes model). *We consider a stationary marked Hawkes process where the conditional intensity $\lambda(t)$ at time $t$ is given by [2]:*

$$\lambda(t) = \mu + \sum_{t_i < t} m_i \phi(t - t_i) \tag{1}$$

*where:*

- $\mu > 0$ *is the constant background (immigration) rate.*
- $\{t_i\}$ *are the event times.*
- $\{m_i\}$ *are i.i.d. marks with* $E[M] = \mu_M > 0$ *and* $E[M^2] < \infty$.
- $\phi(t) \geq 0$ *is the excitation kernel with total integral* $\kappa = \int_0^\infty \phi(t)\,dt < \infty$. *We assume the effective branching ratio* $n := \kappa E[M]$ *satisfies* $0 < n < 1$ *(stationarity).*

**Remark 1** (Unpredictable marks)**.** *Every mark is an i.i.d. copy of $M$, drawn independently of the process history, and enters the dynamics only by scaling the triggering kernel: conditional on an event carrying mark $m$, its offspring form a Poisson process of intensity* $m\,\phi(\cdot)$ (1)*. Offspring marks are independent of the parent's, and immigrant marks share the distribution of $M$; hence for an immigrant mark $M_0$,* $E[M_0] = E[M]$ *and* $E[M_0^2] = E[M^2]$.

**Remark 2** (Physical Interpretation)**.** *In the context of accelerometry, $m_i$ represents the intensity (counts or MIMS units) of the $i$-th active epoch. The i.i.d. mark assumption corresponds to treating intensity as an exogenous attribute of each active epoch, while temporal clustering arises from the self-excitation mechanism encoded in $\phi(t)$.*

**Remark 3** (Stationarity and Real Data)**.** *This derivation assumes a stationary process; non-stationarity in real activity data is addressed in the main text.*

##### 2.2 The Dispersion Index

**Definition 1** (Dispersion Index)**.** *For the sum of marks $S_W = \sum_{i \in W} m_i$ in a window of size $W$, the dispersion index is:*

$$D_{marked} = \frac{Var(S_W)}{E[S_W]} \tag{2}$$

For an *unmarked* Poisson counting process, the analogous dispersion index for counts satisfies $\mathrm{Var}(N_W)/E[N_W] = 1$. In contrast, by Campbell's theorem [3], for a stationary marked Poisson process with background rate $\mu$ over a window of size $W$, the mean and variance of the mark sum $S_W$ are $E[S_W] = \mu W E[M]$ and $\mathrm{Var}(S_W) = \mu W E[M^2]$, giving $D_{\text{marked}} = E[M^2]/E[M] = \mathcal{Q}$. For self-exciting processes, temporal clustering further inflates dispersion beyond this mark-driven baseline.

#### 3 Main Derivation: Cluster Representation

##### 3.1 The Cluster Decomposition

The Hawkes process admits a cluster representation [4]: it can be viewed as a superposition of independent clusters, each initiated by an "immigrant" event arriving according to a Poisson process with rate $\mu$.

Let $K$ denote the number of immigrant events in a window of size $W$. Then $K \sim \mathrm{Poisson}(\mu W)$. Each immigrant generates a cluster consisting of itself plus all its "descendants" (offspring,

offspring of offspring, etc.). Let $C_m$ denote the total mass (sum of marks) of a single cluster. The total marked sum in the window is:

$$S_W = \sum_{k=1}^{K} C_{m,k} \tag{3}$$

where $C_{m,k}$ are i.i.d. copies of $C_m$.

#### 3.2 Compound Poisson Moments

In the large-window regime (or under negligible boundary truncation) where clusters initiated within the window are effectively observed in full, $S_W$ is well-approximated by a compound Poisson sum with i.i.d. cluster masses. Since $S_W$ follows a compound Poisson distribution, standard results give:

$$E[S_W] = E[K] \cdot E[C_m] = \mu W \cdot E[C_m] \tag{4}$$

$$\mathrm{Var}(S_W) = E[K] \cdot E[C_m^2] = \mu W \cdot E[C_m^2] \tag{5}$$

Substituting into Equation 2:

$$D_{\text{marked}} = \frac{\mu W \cdot E[C_m^2]}{\mu W \cdot E[C_m]} = \frac{E[C_m^2]}{E[C_m]} \tag{6}$$

Thus, the dispersion index reduces to the ratio of the second to first moment of the cluster mass. Our task is to calculate these moments.

### 4 Cluster Mass Moments

#### 4.1 Recursive Cluster Structure

**Definition 2** (Cluster Mass). *A cluster initiated by an immigrant with mark $M_0$ consists of the immigrant itself plus all sub-clusters generated by its direct offspring:*

$$C_m = M_0 + \sum_{j=1}^{Z} C_{m,j} \tag{7}$$

*where:*

- *$M_0$ is the mark of the immigrant (a random variable with distribution $M$)*
- *$Z$ is the number of direct offspring generated by this immigrant*
- *$C_{m,j}$ are i.i.d. copies of $C_m$ (masses of sub-clusters initiated by each offspring)*

#### 4.2 The Offspring Distribution

**Lemma 1** (Expected Offspring)**.** *For the marked Hawkes process (Equation 1), an event with mark $M_0$ occurring at time $t_0$ contributes an intensity $M_0\phi(t-t_0)$ to the conditional intensity at all future times $t > t_0$. The expected number of direct offspring generated by this event is:*

$$E[Z|M_0] = \int_{t_0}^{\infty} M_0\phi(t-t_0)\,dt = M_0\int_0^{\infty}\phi(\tau)\,d\tau = M_0\kappa \tag{8}$$

*Proof.* The intensity contribution from the event at $t_0$ is $M_0\phi(t-t_0)$ for all $t > t_0$. Under the cluster representation, each infinitesimal interval $(t, t+dt)$ has probability $M_0\phi(t-t_0)dt$ of producing an offspring event. Integrating this intensity over all future time gives the expected number of events it generates:

$$E[\text{offspring}] = \int_{t_0}^{\infty} M_0\phi(t-t_0)\,dt = M_0\int_0^{\infty}\phi(\tau)\,d\tau = M_0\kappa.$$

□

**Definition 3** (Branching Ratio)**.** *The effective branching ratio $n$ is the expected number of offspring for a generic event (unconditioning over the mark distribution):*

$$n = E[Z] = E_M[E[Z|M_0]] = E_M[M_0\kappa] = \kappa E[M] \tag{9}$$

#### 4.3 First Moment: $E[C_m]$

**Proposition 1** (Expected Cluster Mass)**.** *The expected total mass of a cluster is:*

$$E[C_m] = \frac{E[M]}{1-n} \tag{10}$$

*Proof.* Taking expectations in Equation 7:

$$E[C_m] = E[M_0] + E\left[\sum_{j=1}^{Z} C_{m,j}\right] \tag{11}$$

Using Wald's identity for random sums ($E[\sum_{j=1}^{Z} X_j] = E[Z]E[X]$ when $\{X_j\}$ are i.i.d. and independent of $Z$):

$$E[C_m] = E[M] + E[Z]\cdot E[C_m] \tag{12}$$

Substituting $E[Z] = n$ from Equation 9:

$$E[C_m] = E[M] + n\cdot E[C_m] \tag{13}$$

Solving for $E[C_m]$:

$$E[C_m](1-n) = E[M] \quad\Rightarrow\quad E[C_m] = \frac{E[M]}{1-n} \tag{14}$$

□

#### 4.4 Second Moment: $E[C_m^2]$

**Proposition 2** (Second Moment of Cluster Mass)**.** *The second moment of the cluster mass is:*

$$E[C_m^2] = \frac{E[M^2]}{(1-n)^3} \tag{15}$$

*Proof.* We square Equation 7:

$$C_m^2 = M_0^2 + 2M_0 \sum_{j=1}^{Z} C_{m,j} + \left( \sum_{j=1}^{Z} C_{m,j} \right)^2 \tag{16}$$

We take expectations term by term.

**Term 1:** $E[M_0^2] = E[M^2]$

**Term 2: The Cross Term**

$$\begin{aligned} E\left[2M_0 \sum_{j=1}^{Z} C_{m,j}\right] &= 2E\left[E\left[M_0 \sum_{j=1}^{Z} C_{m,j} \,\middle|\, M_0, Z\right]\right] \\ &= 2E\left[M_0 \cdot Z \cdot E[C_m]\right] \quad \text{(sub-clusters indep. of } M_0, Z) \\ &= 2E[C_m] \cdot E[M_0 \cdot Z] \end{aligned} \tag{17}$$

To evaluate $E[M_0 \cdot Z]$, we use the law of iterated expectations:

$$\begin{aligned} E[M_0 \cdot Z] &= E[M_0 \cdot E[Z|M_0]] \quad \text{(tower property)} \\ &= E[M_0 \cdot \kappa M_0] \quad \text{(from Lemma 1)} \\ &= \kappa E[M_0^2] = \kappa E[M^2] \end{aligned} \tag{18}$$

Combining Equations 17 and 18:

$$\text{Term 2} = 2\kappa E[M^2] E[C_m] \tag{19}$$

**Term 3: The Squared Sum**

For a random sum of i.i.d. variables $X_j$, we use the standard second-moment identity, equivalently the Blackwell–Girshick variance formula:

$$E\left[\left(\sum_{j=1}^{Z} X_j\right)^2 \,\middle|\, Z\right] = Z \cdot E[X^2] + Z(Z-1) \cdot (E[X])^2 \tag{20}$$

This can be verified by expanding $(\sum X_j)^2 = \sum X_j^2 + \sum_{i \neq j} X_i X_j$ and using independence.

Applying this to our case with $X = C_m$:

$$E\left[\left(\sum_{j=1}^{Z} C_{m,j}\right)^2\right] = E\left[Z \cdot E[C_m^2] + Z(Z-1) \cdot (E[C_m])^2\right]$$
$$= E[Z] \cdot E[C_m^2] + E[Z(Z-1)] \cdot (E[C_m])^2 \tag{21}$$

We already know $E[Z] = n$. For the second term, we need $E[Z(Z-1)]$.
For a Poisson random variable with mean $\lambda$, we have $\mathrm{Var}(Z) = \lambda$ and:

$$E[Z^2] = \mathrm{Var}(Z) + (E[Z])^2 = \lambda + \lambda^2 \tag{22}$$

Therefore:

$$E[Z(Z-1)] = E[Z^2] - E[Z] = \lambda + \lambda^2 - \lambda = \lambda^2 \tag{23}$$

In our marked process, $Z|M_0 \sim \mathrm{Poisson}(\kappa M_0)$, so:

$$E[Z(Z-1)|M_0] = (\kappa M_0)^2 \tag{24}$$

Unconditioning over $M_0$:

$$E[Z(Z-1)] = E[(\kappa M_0)^2] = \kappa^2 E[M^2] \tag{25}$$

Substituting into Equation 21:

$$\text{Term 3} = n \cdot E[C_m^2] + \kappa^2 E[M^2] \cdot (E[C_m])^2 \tag{26}$$

**Combining All Terms**

From Equation 16, we have:

$$E[C_m^2] = E[M^2] + 2\kappa E[M^2]E[C_m] + nE[C_m^2] + \kappa^2 E[M^2](E[C_m])^2 \tag{27}$$

Collecting $E[C_m^2]$ terms on the left:

$$E[C_m^2](1-n) = E[M^2]\left[1 + 2\kappa E[C_m] + \kappa^2 (E[C_m])^2\right] \tag{28}$$

The bracketed term is a perfect square:

$$E[C_m^2](1-n) = E[M^2]\left(1 + \kappa E[C_m]\right)^2 \tag{29}$$

Now substitute $E[C_m] = \frac{E[M]}{1-n}$ from Proposition 1:

$$\begin{aligned} 1 + \kappa E[C_m] &= 1 + \kappa \cdot \frac{E[M]}{1-n} = 1 + \frac{\kappa E[M]}{1-n} \\ &= 1 + \frac{n}{1-n} \quad \text{(using } n = \kappa E[M] \text{ from Eq. 9)} \\ &= \frac{1}{1-n} \end{aligned} \tag{30}$$

Substituting Equation 30 into Equation 29:

$$E[C_m^2](1-n) = E[M^2]\left(\frac{1}{1-n}\right)^2 = \frac{E[M^2]}{(1-n)^2} \tag{31}$$

Dividing both sides by $(1-n)$:

$$E[C_m^2] = \frac{E[M^2]}{(1-n)^3} \tag{32}$$

□

### 5 The Final Result

**Theorem 1** (Marked Hawkes Dispersion Index). *For a stationary marked Hawkes process satisfying Assumption 1, the dispersion index in the limit of large windows is:*

$$D_{marked} = \mathcal{Q} \cdot \frac{1}{(1-n)^2} \tag{33}$$

*where the mark-moment ratio is defined as:*

$$\mathcal{Q} = \frac{E[M^2]}{E[M]} \tag{34}$$

*Proof.* Substituting Propositions 1 and 2 into Equation 6:

$$D_{\text{marked}} = \frac{E[C_m^2]}{E[C_m]} = \frac{\frac{E[M^2]}{(1-n)^3}}{\frac{E[M]}{1-n}} = \frac{E[M^2]}{E[M]} \cdot \frac{1}{(1-n)^2} = \mathcal{Q} \cdot \frac{1}{(1-n)^2}$$

□

**Corollary 1** (Branching Ratio Estimator). *Inverting Equation 33 yields the branching ratio estimator:*

$$\boxed{\hat{n} = 1 - \sqrt{\frac{\mathcal{Q}}{D_{marked}}}} \tag{35}$$

### 6 Special Cases and Interpretation

#### 6.1 The Unmarked Case

**Corollary 2** (Unmarked Hawkes Process)**.** *When all events have identical marks ($m_i = c$ for all $i$), we have $E[M] = c$ and $E[M^2] = c^2$, giving $\mathcal{Q} = c$. Equation 33 reduces to:*

$$D_{unmarked} = \frac{c}{(1-n)^2} \tag{36}$$

*Note that here $D_{unmarked}$ is the dispersion of the* marked sum *(i.e., total mass), not the event count. For $c = 1$ (binary events), this recovers the Hardiman & Bouchaud result:*

$$D = \frac{1}{(1-n)^2}, \quad \hat{n} = 1 - \frac{1}{\sqrt{D}} \tag{37}$$

#### 6.2 The compound Poisson process ($n = 0$)

**Corollary 3** (Marked Poisson process)**.** *When there is no self-excitation ($n = 0$), each cluster degenerates to its immigrant alone, $C_m = M_0$, and the process reduces to a stationary marked Poisson process with rate $\mu$ and i.i.d. marks. The windowed mark sum is then a compound Poisson random variable,*

$$S_W = \sum_{i=1}^{N_W} M_i, \qquad N_W \sim \text{Poisson}(\mu W), \tag{38}$$

*with $M_i$ i.i.d. and independent of $N_W$. Its mean and variance are the standard compound Poisson moments*

$$E[S_W] = \mu W\, E[M], \qquad \text{Var}(S_W) = \mu W\, E[M^2], \tag{39}$$

*so the dispersion index is the compound Poisson variance-to-mean ratio*

$$D_{marked}\big|_{n=0} = \frac{\text{Var}(S_W)}{E[S_W]} = \frac{E[M^2]}{E[M]} = \mathcal{Q}. \tag{40}$$

*The compound Poisson structure used at the cluster level for $n > 0$ here collapses to its single-event form, which is the classical compound Poisson moment identity for a marked Poisson process (equivalently, Campbell's theorem [3]).*

#### 6.3 Interpretation of $\mathcal{Q}$

The mark-moment ratio $\mathcal{Q} = E[M^2]/E[M]$ quantifies the variability of marks. Physically, $\mathcal{Q}$ represents the dispersion that would be observed in a marked Poisson process ($n = 0$, no clustering). The factor partitions the observed dispersion into:

$$D_{\text{marked}} = \underbrace{\mathcal{Q}}_{\substack{\text{compound Poisson}\\ \text{baseline}}} \times \underbrace{\frac{1}{(1-n)^2}}_{\text{clustering inflation}} = \underbrace{\mathcal{Q}}_{\text{mark variance}} \times \underbrace{\frac{1}{(1-n)^2}}_{\text{temporal clustering}} . \tag{41}$$

Without dividing by $\mathcal{Q}$, high intensity variance would be misinterpreted as strong temporal clustering.

### References


[1] Stephen J. *Hardiman* and Jean-Philippe *Bouchaud*. "Branching-ratio approximation for the self-exciting Hawkes process". en. In: *Physical Review E* 90.6 (Dec. 2014), p. 062807. ISSN: 1539-3755, 1550-2376. DOI: 10.1103/PhysRevE.90.062807.

[2] Alan G. *Hawkes*. "Spectra of some self-exciting and mutually exciting point processes". en. In: *Biometrika* 58.1 (1971), pp. 83–90. ISSN: 0006-3444, 1464-3510. DOI: 10.1093/biomet/58.1.83.

[3] Daryl J. *Daley* and D. *Vere-Jones*. *An introduction to the theory of point processes*. 2nd ed. New York: Springer, 2003. ISBN: 978-0-387-95541-4 978-0-387-21337-8 978-0-387-49835-5.

[4] Alan G. *Hawkes* and David *Oakes*. "A cluster process representation of a self-exciting process". en. In: *Journal of Applied Probability* 11.3 (Sept. 1974), pp. 493–503. ISSN: 0021-9002, 1475-6072. DOI: 10.2307/3212693.
