## Supplementary material for "Scalable estimation of temporal clustering in accelerometry: a kernel-independent dispersion index grounded in the Hawkes process": S2

#### 1 Model fit and residual diagnostics

The capability of the Hawkes process in capturing the dynamics of active bursts was assessed using a residual analysis framework grounded in the random-time-change theorem [1]. Under correct model specification, the compensator-transformed inter-arrival times $\tau_i^* = \int_{t_{i-1}}^{t_i} \lambda(s)\,\mathrm{d}s = \Lambda(t_i) - \Lambda(t_{i-1})$ constitute independent and identically distributed standard exponential random variables. Departures from this characterisation indicate model misspecification. Three complementary diagnostics were applied to assess goodness of fit.

**Q–Q plots.** Quantile-quantile plots were constructed by plotting the empirical quantiles of $\{\tau_i^*\}$ against the corresponding theoretical quantiles of the Exp(1) distribution. Systematic departures from the 45° reference line indicate regions of misfit, such as overdispersion, underdispersion, or residual clustering.

**Kolmogorov–Smirnov test.** A one-sample Kolmogorov–Smirnov (KS) test was applied to $\{\tau_i^*\}$ against the Exp(1) distribution. The test statistic $D = \sup_{x\geq 0} \left|F_n(x) - (1 - e^{-x})\right|$ measures the maximum pointwise deviation between the empirical cumulative distribution function $F_n$ and the theoretical Exp(1) counterpart, with small $p$-values providing evidence against distributional adequacy.

**Ljung–Box test.** Residual serial dependence was examined using the Ljung–Box portmanteau test applied to $\{\tau_i^*\}$. Failure to reject the null hypothesis of serial independence ($p > 0.05$) indicates that the fitted model has adequately absorbed the autocorrelation structure present in the observed event sequence.

### 2 Study population

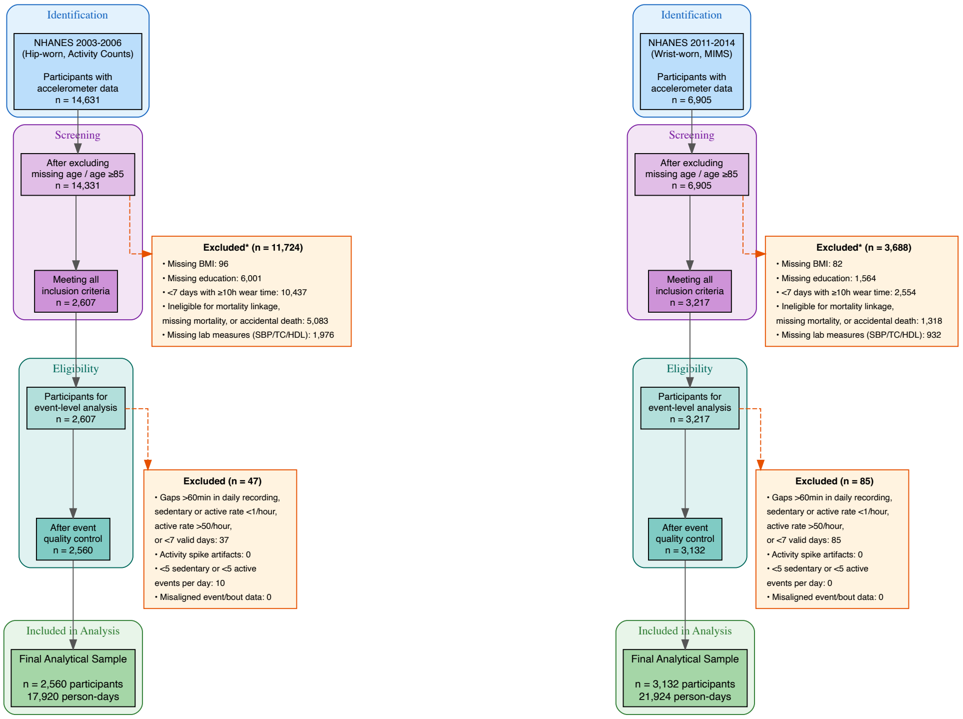


(a) Hip-worn cohort (2003-2006)  (b) Wrist-worn cohort (2011-2014)

Figure 1: Participant flow diagrams.

Table 1: Baseline characteristics of study participants by accelerometer cohort.

| **Characteristic** | **Hip-worn (N=2,560)** | **Wrist-worn (N=3,132)** |
|---|---|---|
| Age, years | 56 (17) | 53 (17) |
| *Gender* | | |
| Male | 1,316 (51%) | 1,386 (44%) |
| Female | 1,244 (49%) | 1,746 (56%) |
| *Racial and ethnic groups* | | |
| White | 1,530 (60%) | 1,348 (43%) |
| Black | 409 (16%) | 710 (23%) |
| Mexican American | 463 (18%) | 350 (11%) |
| Other Hispanic | 46 (1.8%) | 296 (9.5%) |
| Other | 112 (4.4%) | 428 (14%) |
| *Education* | | |
| Less than high school | 619 (24%) | 669 (21%) |
| High school | 619 (24%) | 648 (21%) |
| More than high school | 1,322 (52%) | 1,815 (58%) |
| BMI, $kg/m^2$ | 27.9 (5.5) | 29 (7) |
| *Smoking Status* | | |
| Never | 1,291 (50%) | 1,804 (58%) |
| Former | 843 (33%) | 794 (25%) |
| Current | 426 (17%) | 533 (17%) |
| *Alcohol Consumption* | | |
| Moderate Drinker | 1,381 (54%) | 1,769 (56%) |
| Non-Drinker | 943 (37%) | 1,032 (33%) |
| Heavy Drinker | 150 (5.9%) | 170 (5.4%) |
| Missing | 86 (3.4%) | 161 (5.1%) |
| Systolic BP, mmHg | 127 (20) | 125 (19) |
| *Comorbidities* | | |
| Congestive Heart Failure | 100 (3.9%) | 107 (3.4%) |
| Coronary Heart Disease | 149 (5.8%) | 145 (4.6%) |
| Cancer | 306 (12%) | 360 (11%) |
| Stroke | 85 (3.3%) | 125 (4.0%) |
| Diabetes | 285 (11%) | 476 (15%) |
| *Mobility Problem* | | |
| No Difficulty | 2,050 (80%) | 2,441 (78%) |
| Any Difficulty | 510 (20%) | 691 (22%) |
| *Activity Metrics* | | |
| Wear Time, min/day | 912 (133) | 1,369 (57) |
| Total Activity Count | 250,304 (127,836) | 12,639 (3,394) |
| MVPA, min/day | 20 (21) | 258 (109) |
| Peak 30-min Activity | 2,199 (1,046) | 41 (9) |
| ASTP | 0.29 (0.08) | 0.23 (0.06) |
| Sedentary Time, min/day | 1,082 (106) | 957 (123) |
| Sedentary Bout Duration, min | 12.7 (3.8) | 9.98 (2.40) |
| Active Bout Duration, min | 3.94 (1.23) | 4.99 (1.61) |
| *Sleep & time-of-day (TOD) pattern (days, 0–7)* | | |
| Sleep Consolidation (CoV) | † | 1.12 (0.29) |
| Consolidated Sleep, median (IQR) | † | 0 (0–1) |
| Days with Early Activity, median (IQR) | † | 5 (3–6) |
| Days with Late Activity, median (IQR) | † | 2 (0–4) |

Values are mean (SD), n (%), or median (IQR). ASTP = Active-to-Sedentary Transition Probability. CoV = Coefficient of Variation. Early/late activity and consolidated sleep range from 0 to 7 days.
† Unavailable for the hip cohort; participants were instructed to remove the device at bedtime.

### 3 Example individual data

#### 3.1 Data visualisation

These figures supplement the main-text results by providing day-by-day visualizations of the event-extraction pipeline (described under event extraction in the main-text Materials and methods). Supplementary Figures 2–15 show representative participants across all seven monitoring days. Each figure contains three panels: (Top) Raw minute-level accelerometry data showing activity counts (hip) or MIMS units (wrist); (Middle) Event classification into sedentary (blue) and active (green) states based on intensity thresholds (hip: $<100$ counts =

sedentary, $\geq$100 = active; wrist: <10.558 MIMS = sedentary, $\geq$10.558 = active); (Bottom) Marked Hawkes events showing the extracted active epochs with their intensity marks (later normalized to mean = 1 per day). These visualizations demonstrate the within-person consistency of TOD patterns and the conversion from continuous time series to continuous point process events suitable for Hawkes modeling. The clustering structure visible in these figures—activity epochs occurring in temporal clusters rather than uniformly—is quantified by the dispersion index and branching ratio metrics reported in the main text.

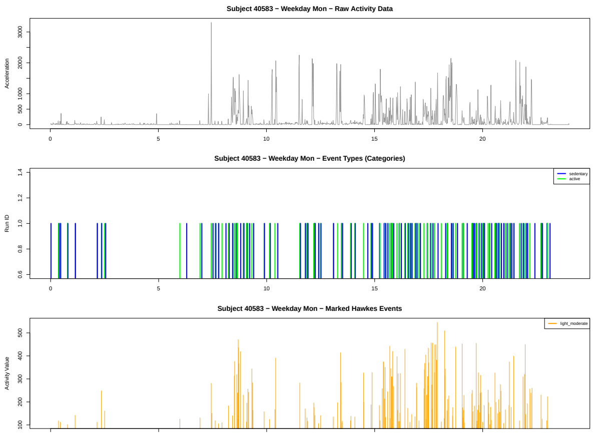


Figure 2: Hip-worn cohort (Subject 40583) – Monday.

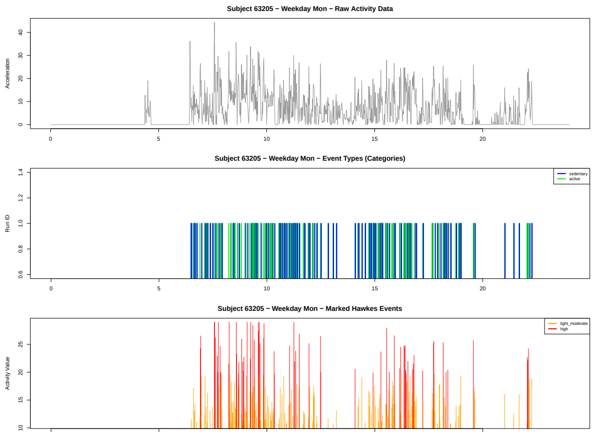


Figure 3: Wrist-worn cohort (Subject 63205) – Monday.

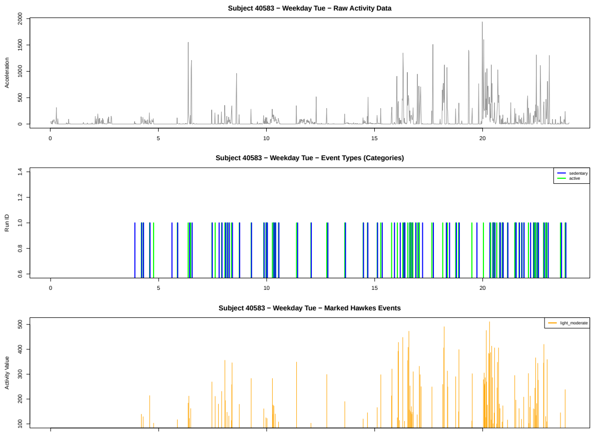


Figure 4: Hip-worn cohort (Subject 40583) – Tuesday.

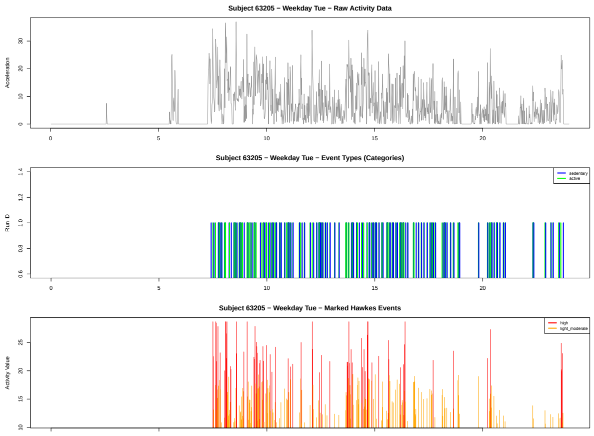


Figure 5: Wrist-worn cohort (Subject 63205) – Tuesday.

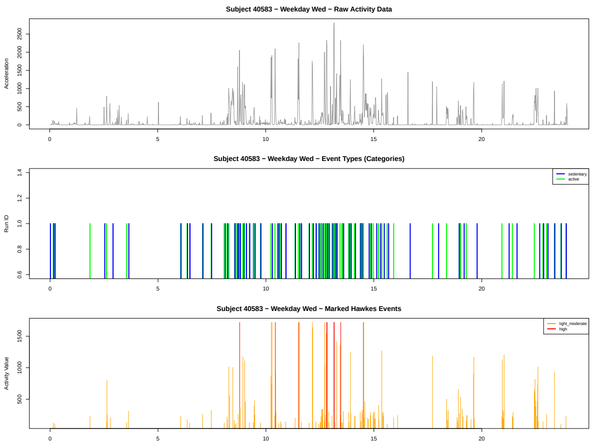


Figure 6: Hip-worn cohort (Subject 40583) – Wednesday.

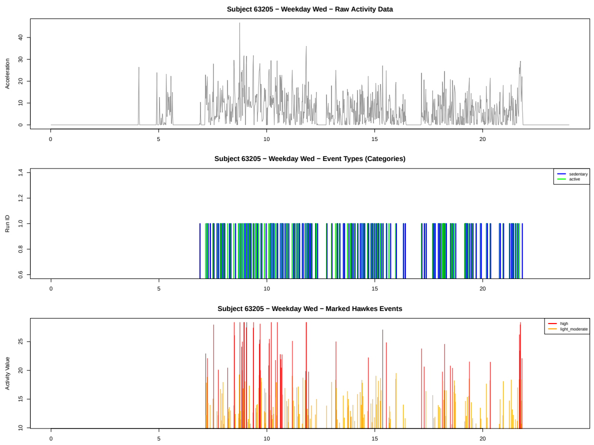


Figure 7: Wrist-worn cohort (Subject 63205) – Wednesday.

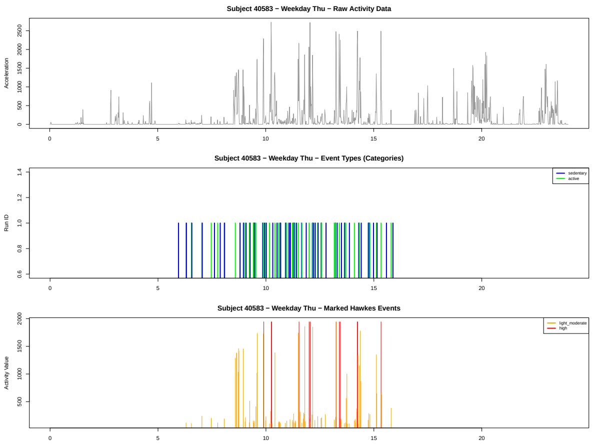


Figure 8: Hip-worn cohort (Subject 40583) – Thursday.

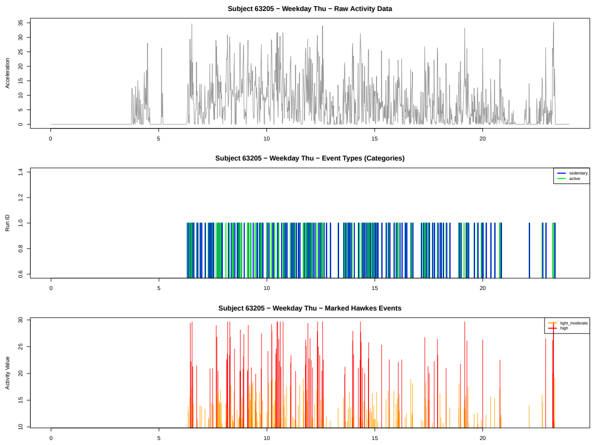


Figure 9: Wrist-worn cohort (Subject 63205) – Thursday.

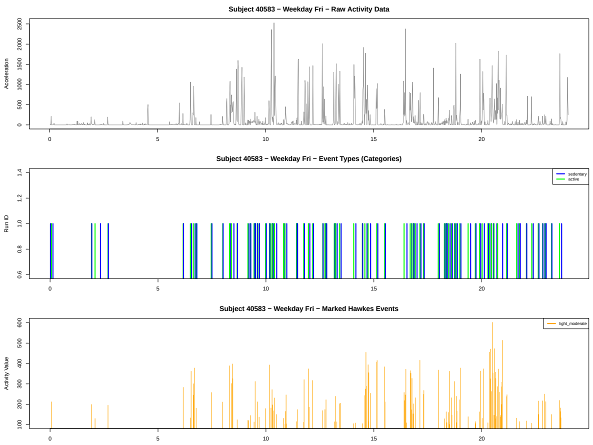


Figure 10: Hip-worn cohort (Subject 40583) – Friday.

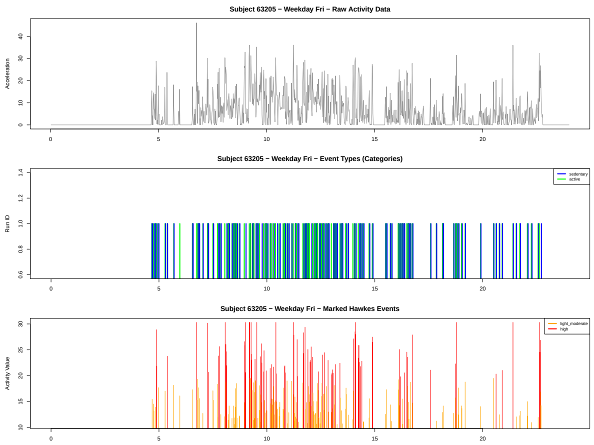


Figure 11: Wrist-worn cohort (Subject 63205) – Friday.

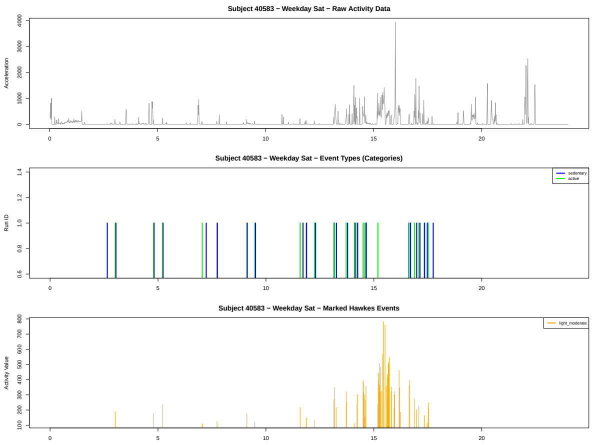


Figure 12: Hip-worn cohort (Subject 40583) – Saturday.

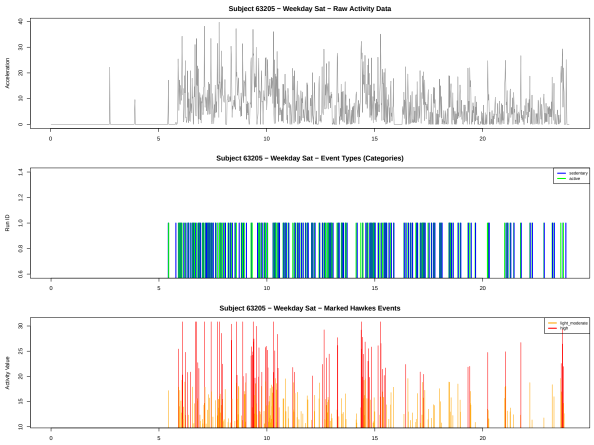


Figure 13: Wrist-worn cohort (Subject 63205) – Saturday.

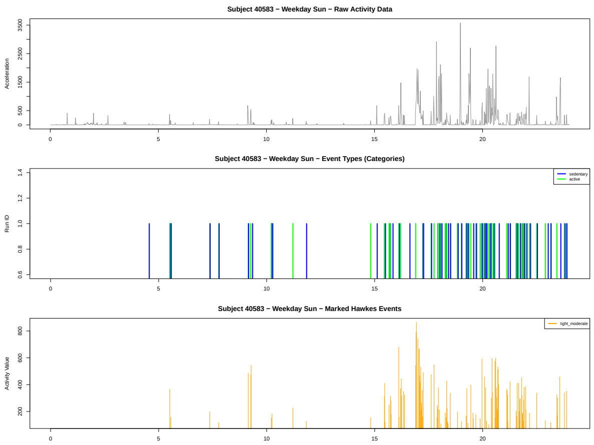


Figure 14: Hip-worn cohort (Subject 40583) – Sunday.

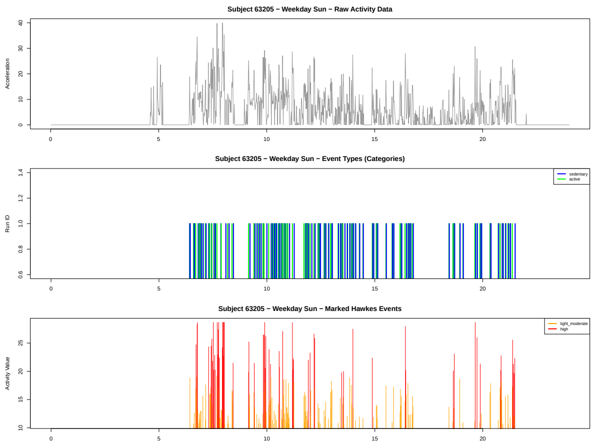


Figure 15: Wrist-worn cohort (Subject 63205) – Sunday.

#### 3.2 Goodness-of-fit evaluation of unmarked and marked models

Supplementary Figures 16 to 19 show goodness-of-fit diagnostics for the unmarked and marked Hawkes models: conditional intensity trajectories, KS D-statistic, Q-Q plots against Exp(1), and inter-arrival time patterns. Ljung–Box tests confirmed roughly no significant residual autocorrelation.

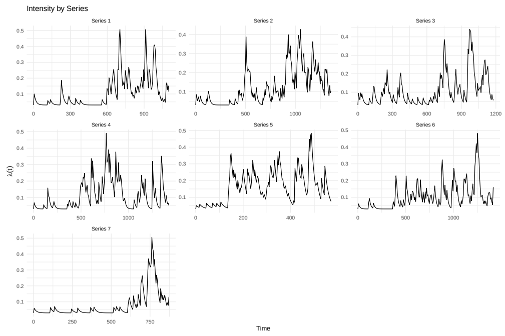


(a) Marked model: conditional intensity

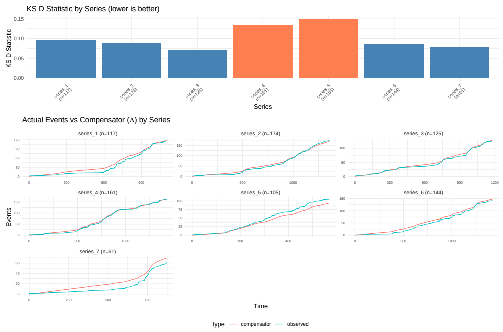


(b) Marked model: KS test

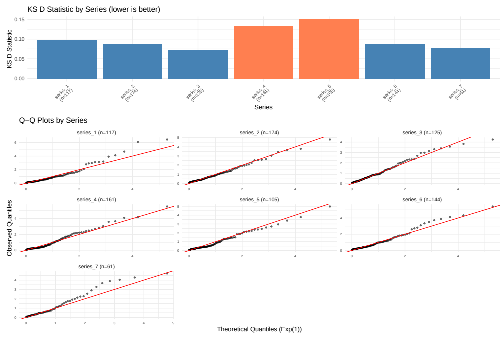


(c) Marked model: Q-Q plots

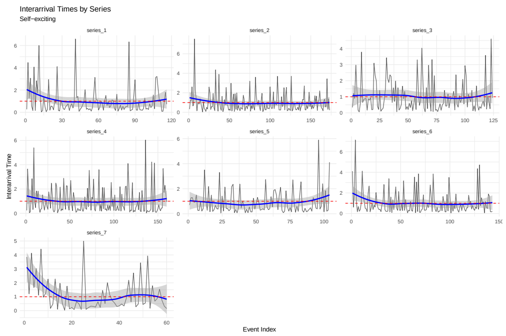


(d) Marked model: inter-arrival pattern

Figure 16: Goodness-of-fit diagnostics for hip-worn cohort MLE models (marked).

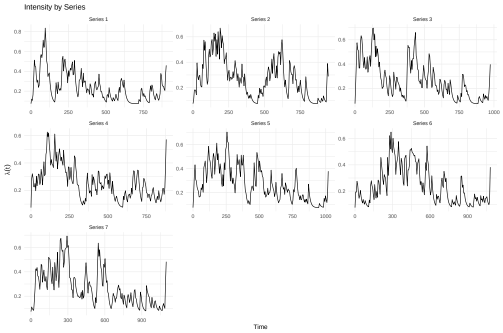


(a) Marked model: conditional intensity

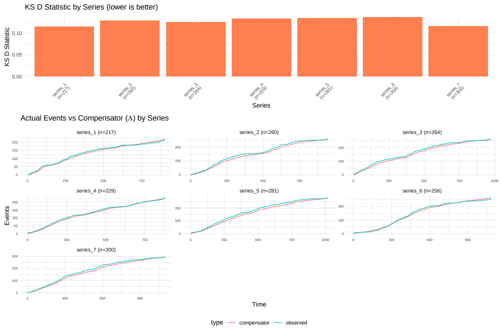


(b) Marked model: KS test

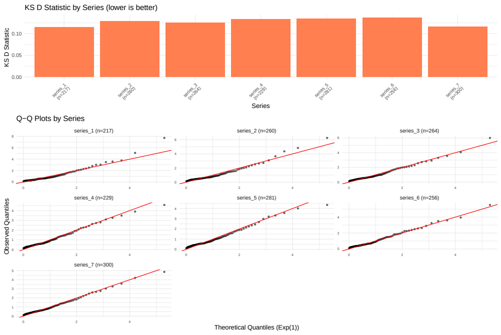


(c) Marked model: Q-Q plots

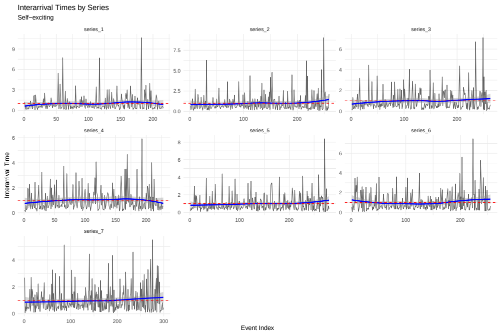


(d) Marked model: inter-arrival pattern

Figure 17: Goodness-of-fit diagnostics for wrist-worn cohort MLE models (marked).

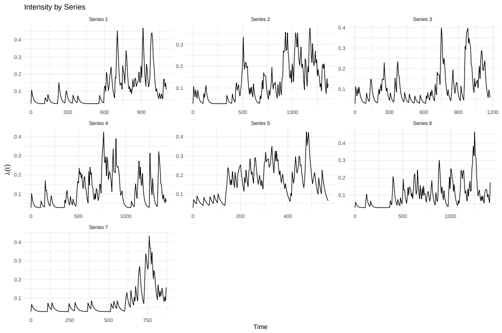


(a) Unmarked model: conditional intensity

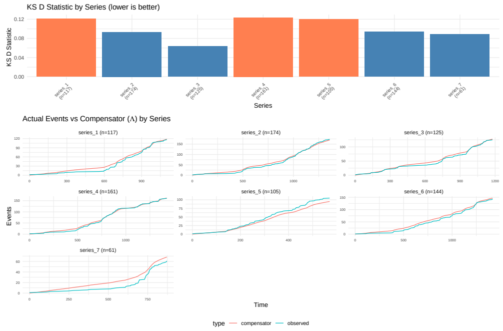


(b) Unmarked model: KS test

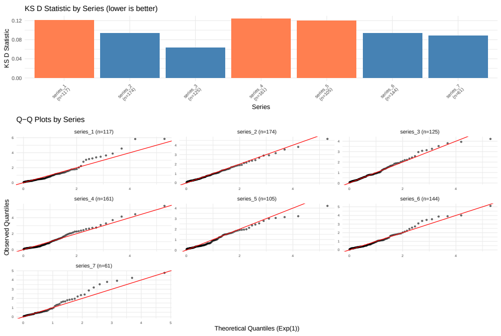


(c) Unmarked model: Q-Q plots

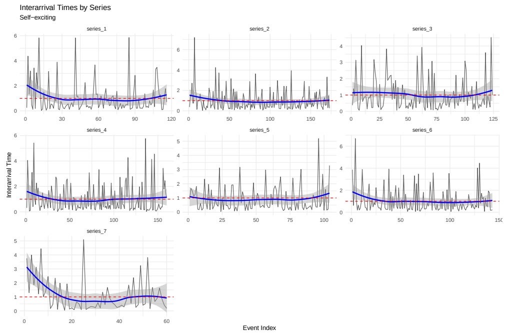


(d) Unmarked model: inter-arrival pattern

Figure 18: Goodness-of-fit diagnostics for hip-worn cohort MLE models (unmarked).

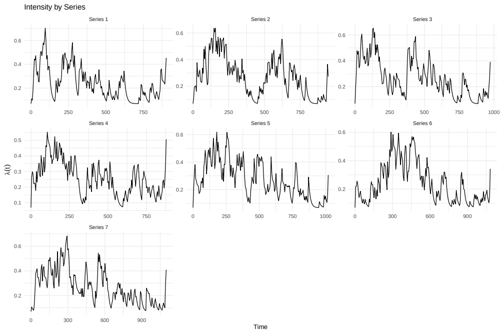


(a) Unmarked model: conditional intensity

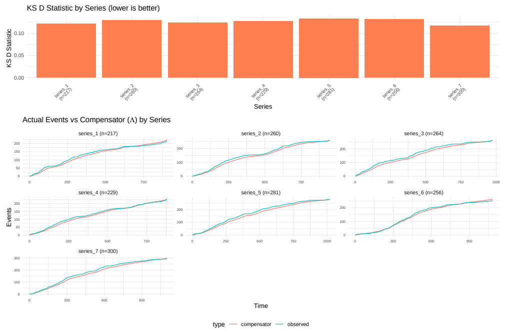


(b) Unmarked model: KS test

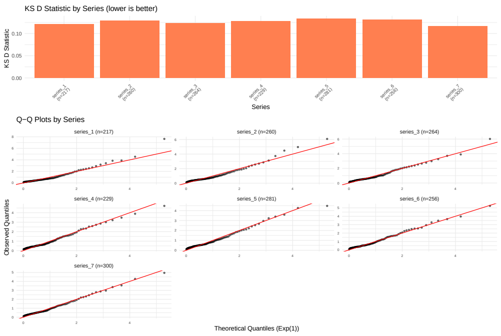


(c) Unmarked model: Q-Q plots

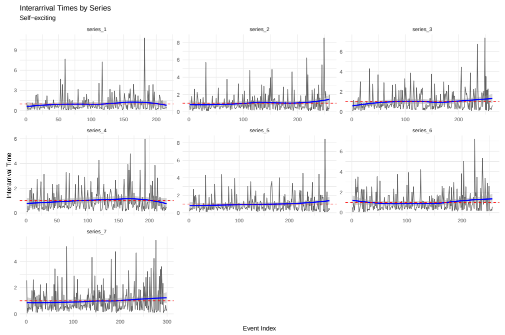


(d) Unmarked model: inter-arrival pattern

Figure 19: Goodness-of-fit diagnostics for wrist-worn cohort MLE models (unmarked).

### 4 Nocturnal activity and all-cause mortality

To characterize nocturnal activity patterns, we derived four variables from accelerometer event data: **1. Sleep consolidation.** For each person-day, we calculated the coefficient of variation (CoV) of activity event counts across four one-hour windows during core sleep time (1-5am). We then averaged person-day CoVs across active nights (those with at least one detected event during 1-5am) per participant; participants for whom all monitored nights had zero detected activity were assigned the maximum observed mean CoV, representing the upper bound of sleep consolidation. Higher CoV indicates activity concentrated in fewer hours (e.g., a single brief awakening), while lower CoV indicates activity distributed across the sleep period (e.g., repeated awakenings or sustained restlessness). **2. Mean nocturnal activity.** For each person-day, we counted activity events during core sleep hours (15am) and averaged these daily counts across the monitoring period. Higher values indicate greater nighttime disturbance, capturing the volume of nocturnal movement independent of its temporal distribution. **3. Early activation.** For each person-day, we calculated the proportion of daily activity events occurring before 8am. Days exceeding 10% were classified as early-active, and we counted the number of such days per participant. Higher values indicate a tendency toward morning-concentrated activity. **4. Late activation.** Analogously, we calculated the proportion of daily activity events occurring after 10pm. Days exceeding 10% were classified as late-active, and we counted the number of such days per participant. Higher values indicate a tendency toward evening-concentrated activity.

Together, sleep consolidation and mean nocturnal activity capture complementary dimensions of nighttime behavior: the former measures *pattern* (fragmented vs. consolidated), while the latter measures *amount* (total nocturnal movement). To explore whether temporal dynamics extend to nighttime behaviour, we examined associations between sleep-related variables and all-cause mortality (Table 2), including sleep consolidation, early activation, and late activation frequencies across the monitoring period.

Table 2: Association between nighttime activity patterns and all-cause mortality in the wrist-worn cohort (2011–2014). HR (95% CI) reported. Adjusted for age, sex, race and ethnicity group.

| **Variable** | **HR (95% CI)** | $p$ |
|---|---|---|
| *Nighttime activity patterns* | | |
| Sleep consolidation (CoV) | 0.645 (0.435–0.955) | 0.029* |
| Mean nocturnal activity | 1.003 (0.995–1.012) | 0.439 |
| Days with early activity | 1.117 (1.036–1.205) | 0.004** |
| Days with late activity | 1.066 (0.993–1.145) | 0.079. |
| *Covariates* | | |
| Age (per year) | 1.117 (1.097–1.136) | <0.001*** |
| Female | 1.015 (0.726–1.420) | 0.929 |
| Race: Mexican American | 0.889 (0.505–1.564) | 0.683 |
| Race: Other Hispanic | 0.637 (0.396–1.026) | 0.064. |
| Race: Black | 0.989 (0.704–1.390) | 0.951 |
| Race: Other | 0.815 (0.470–1.413) | 0.467 |

.$p < 0.1$, *$p < 0.05$, **$p < 0.01$, ***$p < 0.001$. Reference: White, Male. Concordance = 0.85.

Sleep consolidation — days where nocturnal activity was absent or concentrated in fewer hours rather than distributed across the 1–5 am window — was protective: lower CoV in nocturnal activity, indicating more consolidated sleep, was associated with lower mortality (HR = 0.645, 95% CI: 0.435–0.955, $p = 0.029$). This confirmed that distributed nocturnal activity indicative of fragmented sleep is a mortality risk marker, whereas concentrated activity (e.g., a single brief awakening) reflects more restorative rest.

Mean nocturnal activity volume was not associated with mortality, indicating that the *pattern* of nighttime activity, rather than its *amount*, carries prognostic information. Days with early activity were notably associated with higher mortality risk (HR = 1.117, 95% CI: 1.036–1.205, $p = 0.004$), which may reflect restless pre-waking arousals or upper-limb micro-movements rather than volitional early physical activity, given the greater sensitivity of wrist accelerometers to distal limb motion during sleep. Days with late activity showed a similar but non-significant trend (HR = 1.066, $p = 0.079$).

CoV used for nighttime consolidation is conceptually related to the dispersion index ($D = \mathrm{CoV}^2 \cdot \mu$) primarily used for daytime activity patterns: both quantify variability relative to mean activity levels. CoV was chosen for nighttime analysis because the fixed nocturnal window provides a natural unit-free comparison across individuals, whereas the dispersion index is suited for continuous-time event data where comparison to Poisson regularity is meaningful. Together, these findings suggest that temporal patterning of activity—whether measured during waking or sleeping hours—carries prognostic information beyond total activity volume.

### 5 Metrics correlation

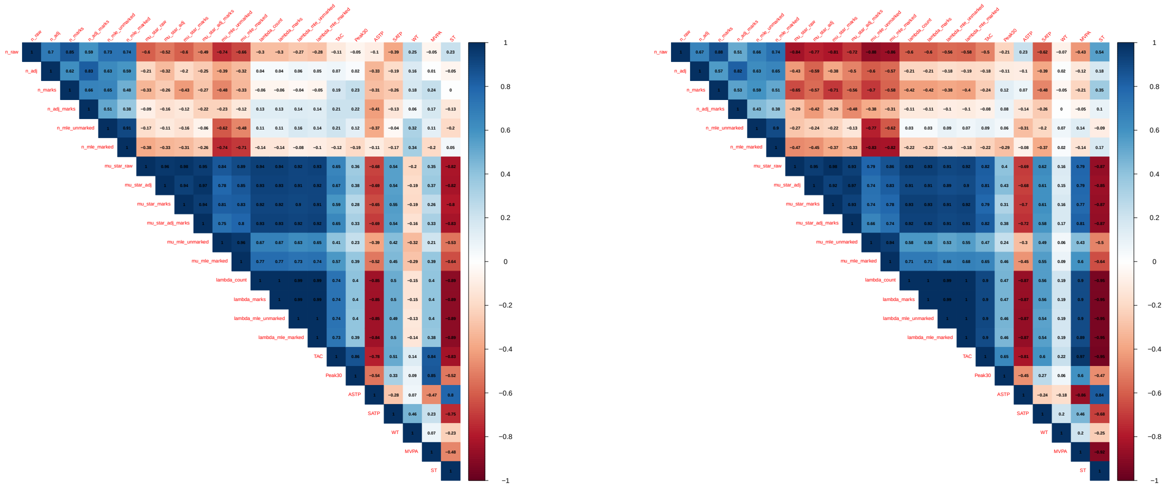

(a) Hip-worn cohort (b) Wrist-worn cohort

Figure 20: Correlation matrices showing relationships between dispersion-based and MLE-estimated Hawkes parameters, and traditional activity metrics (TAC: total activity counts/MIMS; MVPA: minutes of moderate-to-vigorous physical activity; Peak30: mean activity level during the 30 most active minutes of the day; ASTP: active-to-sedentary transition probability; SATP: sedentary-to-active transition probability; ST: sedentary time; WT: wear time).

### References


[1] Emery N. *Brown* et al. "The Time-Rescaling Theorem and Its Application to Neural Spike Train Data Analysis". en. In: *Neural Computation* 14.2 (Feb. 2002), pp. 325–346. ISSN: 0899-7667, 1530-888X. DOI: 10.1162/08997660252741149.
